# Causal Modulation of Brain–Body Habituation Dynamics Implicates Salience Network Mechanisms in Anxiety

**DOI:** 10.64898/2026.01.05.26343461

**Authors:** Jennifer Legon, Yunruo Ni, Wynn Legon

**Affiliations:** Fralin Biomedical Research Institute at Virginia Tech Carilion, Roanoke, VA, 24016, USA; School of Neuroscience, Virginia Polytechnic Institute and State University, Blacksburg, VA, 24061, USA; Graduate Program in Translational Biology, Medicine, and Health, Virginia Polytechnic Institute and State University, Roanoke, VA, 24016, USA; Center for Human Neuroscience Research, Fralin Biomedical Research Institute at Virginia Tech Carilion, Roanoke, VA, 24016, USA; Center for Health Behaviors Research, Fralin Biomedical Research Institute at Virginia Tech Carilion, Roanoke, VA, 24016, USA; Department of Neurosurgery, Carilion Clinic, Roanoke, VA, 24016, USA

**Keywords:** Focused ultrasound, anxiety, neuromodulation, insula, cingulate, autonomic

## Abstract

Anxiety is marked by exaggerated vigilance which may be linked to impaired sensory habituation, yet the physiological mechanisms linking anxiety to dysregulated habituation in the brain remain unclear. The brain’s ability to filter sensory input is characteristic of healthy functioning and disruption of this can lead to over-responsiveness and hypervigilance that may underlie pathological anxiety. Here, we examined how state and trait anxiety relate to acoustic startle reflex habituation across somatic (EMG), cortical (EEG), and autonomic (electrodermal; EDR) responses, and whether casual modulation of the right anterior insula (AI) or anterior mid-cingulate cortex (aMCC) using low-intensity focused ultrasound (LIFU) modulates these dynamics. As part of a larger study, forty participants (median STAI-T = 39.5; range 25 – 68) completed three LIFU sessions (AI, aMCC, Sham) while EMG, EEG, and EDR were recorded during 12 acoustic startle stimuli both before and after LIFU. Habituation slopes were computed separately for early (Trials 2–6) and late (Trials 7–12) windows using Theil–Sen estimators and analyzed with linear mixed-effects models controlling for baseline magnitude (Trial 1) and anxiety covariates.

Before LIFU, higher trait anxiety predicted weaker EMG habituation (ρ ≈ 0.3), whereas state anxiety showed no reliable association with any modality. Across sessions, early slopes were steeper than late slopes for all measures, indicating reduced adaptation with repetition. LIFU to AI or aMCC did not alter EMG or EEG habituation but significantly enhanced early-phase EDR habituation relative to sham, suggesting transient facilitation of autonomic adaptation. Cross-modal analyses revealed robust coupling between EMG and EEG habituation (ρ ≈ 0.4–0.5, p < 0.01), independent of anxiety, whereas EDR habituation varied independently. Mean response magnitudes of EMG, EEG or EDR across trials were unrelated to anxiety and were not affected by LIFU. These findings identify coordinated cortical-somatic habituation as an anxiety-relevant biomarker and demonstrate that the AI and aMCC are involved in autonomic adaptation to sensory stimuli. Neuromodulation of these regions may provide potential therapeutic benefit to reduce autonomic reactivity to anxiety provoking stimuli.

## Introduction

Anxiety disorders are the most common form of mental illness, affecting more than a quarter of the population and representing the sixth leading cause of disability worldwide (1,2). Despite this global burden, more than 75% of individuals with anxiety never receive treatment (3), and among those who do, fewer than half experience substantial improvement (4–6). Given the debilitating and persistent nature of anxiety and the limited efficacy of existing therapies, there is an urgent need for neuroscience-based biomarkers and mechanistic approaches to guide intervention development.

Anxiety can be defined as a sustained emotional state of tension, vigilance, and worry in anticipation of uncertain or potential threat. State anxiety reflects transient feelings of apprehension and physiological arousal in response to environmental demands, whereas trait anxiety represents a stable disposition to perceive threat across contexts. Elevated trait anxiety predisposes individuals to heightened state responses, shaping defensive behavior and physiological reactivity (7,8).

While cognitive and behavioral inhibition have been extensively studied in anxiety (9–11), sensory habituation—how the brain attenuates responses to repetitive stimuli—has received far less attention. Habituation represents a fundamental neural mechanism that protects the brain from sensory overload by reducing responses to persistent or repeated stimuli (12). Impaired habituation has been observed in some psychiatric conditions (13) and may reflect a failure of sensory gating (12). This failure may contribute to exaggerated vigilance and threat sensitivity in anxiety disorders.

Startle paradigms, including the acoustic startle reflex (ASR)(14), pre-pulse inhibition (15), and the auditory P50 potential (15), provide robust tools to probe sensory inhibition. Although findings have been mixed, some studies report diminished or altered mean startle amplitudes in individuals with anxiety and depression (16) though to our knowledge there is no research examining habituation dynamics of this response in the context of anxiety. Notably, the ASR as measured by EMG of the orbicularis oculi muscle shows rapid habituation across the first few trials followed by a slower decline during later trials with each of these phases potentially indexing different sub-cortical reflex loops or cortical habituation machinery. Indeed, several studies in animal and human have implicated brain regions such as the amygdala (12) and cingulate cortex (17) in sensory gating mechanisms – brain areas also implicated in anxiety (18,19). Traditionally, habituation blocks to startle stimuli are suggested (20) and performed to achieve a stable response and avoid confounding subsequent analyses (21), yet they may contain valuable information such as individual differences in habituation dynamics that could underlie state and trait anxiety. Here, we examined whether early and late phases of ASR habituation as measured by EMG predict state and trait anxiety. We also recorded EEG and electrodermal responses (EDR) to the repeated startle stimuli to determine whether cortical and autonomic measures exhibit parallel habituation patterns. We further used low-intensity focused ultrasound (LIFU) (22) to causally test whether modulating the right anterior insula (AI) or anterior mid-cingulate cortex (aMCC) alters these dynamics. We hypothesized that higher anxiety would be associated with reduced sensory inhibition—manifested as flatter early and late-phase habituation slopes—and that LIFU would increase or normalize these slopes, particularly for cortical (EEG) and autonomic (EDR) responses reflecting better habituation. Habituation dynamics to provoking stimuli may provide a functional biomarker of sensory and saliency mechanisms that may help to explain the aberrant increased vigilance to stimuli commonly seen in anxiety disorders.

## METHODS

### Participants

The Virginia Tech Institutional Review Board approved all experimental procedures (VT-IRB #23-192). This study was registered on ClinicalTrials.gov # NCT05839847. N = 40 healthy participants (12M/28F, aged 26.2 ± 5.8 years); range (18–42), who met all inclusion/exclusion criteria and provided written informed consent to all aspects of the study and were financially compensated for participation. Inclusion criteria were females and males ages 18-45. Exclusion criteria were in accordance with contraindications to noninvasive neuromodulation as outlined by Rossi et al. (2009). Exclusions included contradictions for magnetic resonance imaging (MRI) and computed tomography (CT), a history of neurological disorders (e.g., Parkinson’s disease, epilepsy, multiple sclerosis, etc.), head injury resulting in loss of consciousness for >10 minutes, any current treatment or medical disorder with potential nervous system effects, pregnancy, history of alcohol or drug dependence, and/or history of or current cardiovascular disease.

### Overall Study Design

These data were part of a larger study examining the effect of LIFU on anxiety and fear potentiated responses from the NPU threat task(23). This was a single-blind, sham-controlled, cross-over design. Data were collected in four visits over 4 separate days with a minimum of 48 hours between each visit to mitigate any potential carry-over effects. Following informed consent, participants completed the trait anxiety inventory (STAI-T)(7,8) as well as a structural brain MRI and CT scan for the purpose of low-intensity focused ultrasound (LIFU) targeting and acoustic modelling. The remaining three visits were formal testing sessions of LIFU to the right anterior insula (AI), right dorsal anterior cingulate cortex (aMCC), or sham, randomized between and within subjects. At each formal testing session (prior to LIFU), participants completed a review of symptoms (ROS) questionnaire and state anxiety inventory (STAI-S) (7,8). They were then seated in a comfortable chair with arm and neck support and connected to continuous electromyogram (EMG), electroencephalogram (EEG), electrocardiogram (ECG) and electrodermal (EDR) monitoring. A ∼10-minute baseline was collected while the participant rested in the chair. After this, startle habituation was performed (21) and then participants underwent LIFU to either the AI, aMCC, or Sham for 40 seconds (24). The identical startle habituation procedure was conducted roughly 5 minutes after the end of LIFU (**Figure 1A**). In the larger study, then participants performed an NPU threat task (23). This paper will only be describing the results of the startle habituation pre/post LIFU from the EMG, EEG and EDR recordings.

**Figure 1.**
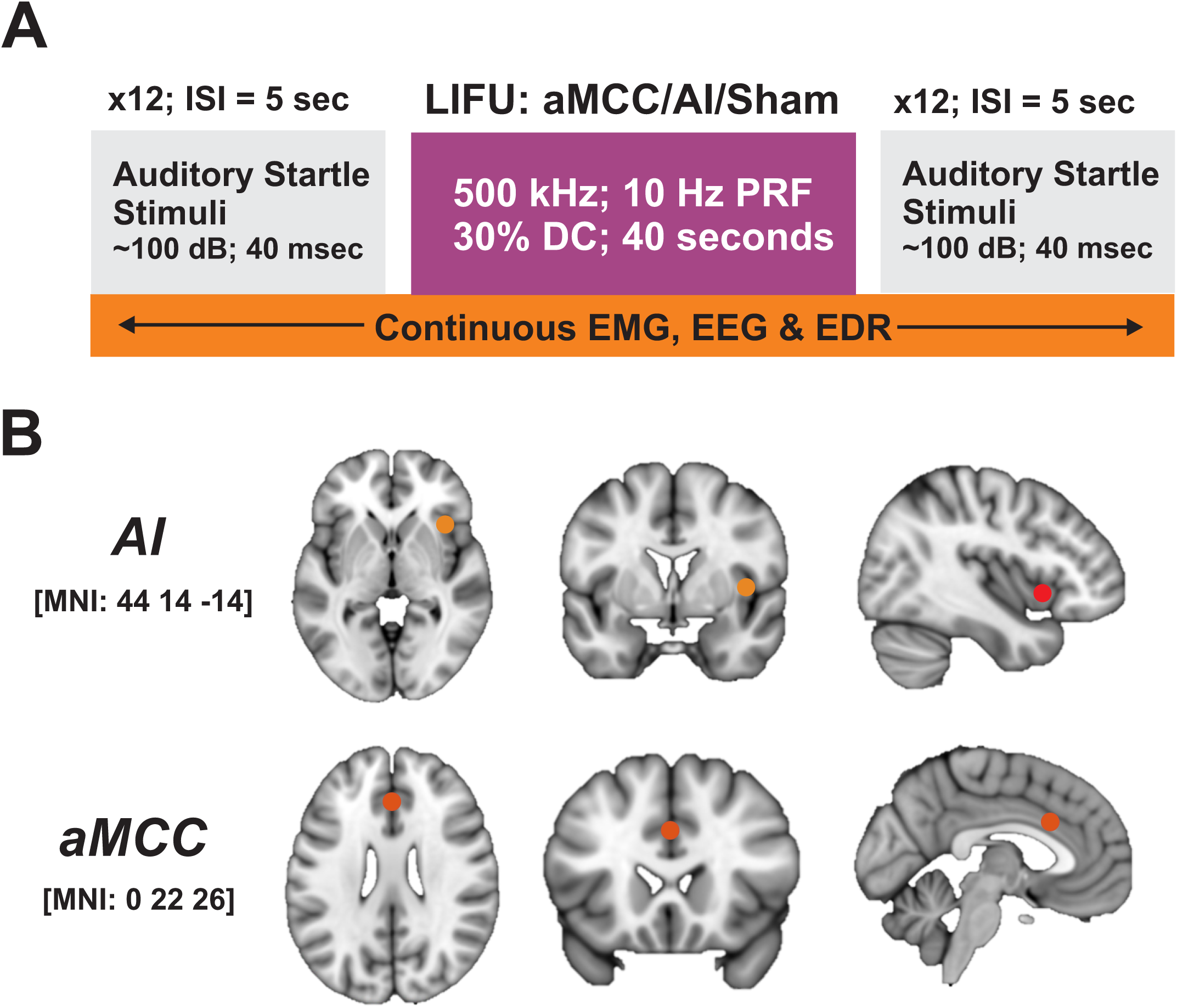
Overall Study Design. **A.** *Experimental timeline*. This study was part of a larger study investigating the effects of LIFU to the right anterior insula (AI), anterior mid-cingulate cortex (aMCC) or Sham on an anxiety/fear task. Prior to the task participants underwent startle habituation that consisted of 12 auditory startle probes (∼100 dB, 40 msec) delivered every 5 seconds. Participants then underwent 40 seconds of LIFU to either the AI, aMCC or Sham on separate days. Roughly 5 minutes after the end of LIFU the same startle habitutation procedure was collected. The response to auditory startle probes were collected using electromyaography (EMG) from the obicularis oculi muscle; electroencephalography (EEG) from vertex scalp channel CZ and using electrodermal responses (EDR) recorded from the hand. **B.** Participants (N = 40) underwent three randomized sessions with LIFU targeted to the right anterior insula (AI), right anterior mid-cingulate cortex (aMCC), or active sham. 5 mm ROIs are placed at MNI coordinate [44 14 -14] for AI targeting and [0 22 26] for aMCC targeting.

#### Startle stimuli

Startle stimuli were square-wave white noise at ∼103 dB for a 40ms duration delivered through over-the-ear headphones. 12 total stimuli were delivered with a constant interstimulus interval of 5 seconds. *Electromyography (EMG).* Two electrodes (Gereonics Inc., miniature silver/silver chloride 405 series) were placed underneath the right eye overlapping the orbicularis oculi muscle. Subjects were instructed to look forward and the first electrode (+) was positioned so that it was placed directly below the pupil. The second electrode (-) was placed 1mm laterally from the first electrode. The wells of the electrodes were filled with a small amount of conductive gel (Spectra 360, Parker Laboratories Inc NJ).

#### Electroencephalography (EEG)

Surface EEG was collected from the vertex channel (CZ) using a 10 mm silver-over-silver chloride cup electrode referenced to the right mastoid. Data were continuously acquired at a 1 kHz sampling rate using a DC amplifier (EEG100C & MP160, Biopac Systems, CA, USA) and AcqKnowledge 5.0 software (BioPac Systems, CA, USA). The scalp was prepared with a mild abrasive gel (Nuprep; Weaver and Company) and rubbing alcohol. Cup electrodes were filled with a conductive paste (Ten20 Conductive; Weaver and Company) and held in place with medical tape. Electrode impedances were verified (<50 kΩ) before recording. Data were stored on a PC for offline data analysis.

#### Electrodermal response (EDR)

Two silver-over silver chloride electrodes were applied to the distal second and third digits the participants’ right hand with isotonic sodium chloride cream (BIOPAC GEL101A) and secured with medical tape. EDR data were sampled at 1 kHz using a DC amplifier (EDA100D & MP160, Biopac Systems, CA, USA) with a constant voltage of 0.5 V and recorded using the AcqKnowledge 5.0 software and stored on a PC for offline data analysis.

#### MRI and CT Imaging

MRI data were acquired on a Siemens 3T Prisma scanner (Siemens Medical Solutions, Erlangen, Germany) at the Fralin Biomedical Research Institute’s Human Neuroimaging Laboratory. Anatomical scans were acquired using a T1-weighted MPRAGE sequence with a TR = 1400 ms, TI = 600 ms, TE = 2.66 ms, flip angle = 12°, voxel size = 0.5x0.5x1.0 mm, FoV read = 245 mm, FoV phase of 87.5%, 192 slices, ascending acquisition.

Computerized Tomography (CT) scans were also collected with a Kernel = Hr60 in the bone window, FoV = 250 mm, kilovolts (kV) = 120, rotation time = 1 second, delay = 2 seconds, pitch = 0.55, caudocranial image acquisition order, 1.0 mm image increments for a total of 121 images and scan time of 13.14 seconds.

#### Electrocardiogram (ECG)

Two latex-free ECG electrodes (MedGel, MDSM611903) were attached symmetrically to the anterior surface of the bilateral forearms immediately distal to the antecubital fossa and grounded to the right elbow. ECG data were continuously collected and sampled at 1 kHz using a DC amplifier (ECG100D & MP160, Biopac Systems, CA, USA) and AcqKnowledge 5.0 software and stored on a PC for offline data analysis.

#### LIFU Transducer

For the AI we used a Sonic Concepts H-281 single-element 500 kHz transducer with an active diameter of 45.0 mm and a geometric focus of 45.0 mm. The focal depth from the exit plane was 38.0 mm. The transducer also had a solid water coupling over the radiating surface to the exit plane. For the aMCC we used a Sonic Concepts H-104 single-element 500kHz transducer with an active diameter of 64mm and a geometric focus of 63.2mm. The focal depth from the exit plane was 52mm.

#### LIFU waveform

LIFU waveforms were generated using a two-channel, 2-MHz function generator (BK 4078B Precision Instruments). Channel 1 was used to gate channel 2 which was a 500 kHz sine wave. Channel 1 was a tapered 5Vp-p square wave burst of 10 Hz (N = 400) with a pulse duration of 30 milliseconds and a pulse repetition interval of 100ms. This resulted in a 40-sec total LIFU application time to each brain region with a duty cycle of 30% (24). The output of channel 2 was sent through a 100-W linear RF amplifier (E&I 2100L; Electronics & Innovation) before being sent to the LIFU transducer. The peak negative pressure of the waveform outside the head was ∼900 kPa or 27 W/cm^2^ spatial peak pulse average intensity (I_SPPA_) for all participants.

#### LIFU targeting

The transducer was coupled to the head using conventional ultrasound gel and custom mineral oil/polymer coupling pucks (25). These pucks have negligible attenuation at 500 kHz and can be made with varying stand-off heights that allow for precise axial (depth) targeting based on individual target depths. Each participant’s right AI and aMCC targets were based on meta-analytic evidence of the convergence of induced and pathological anxiety during unpredictable shock (18) and automated meta-analysis based on the search term ‘anxiety’ (26). The right AI target was Montreal Neurologic Institute (MNI) coordinate [44 14 -14] while the right aMCC target was MNI coordinate [0 22 26] (**Figure 1B**). Depth to each target was measured from the scalp. An appropriate coupling puck was made so that the focal spot of the transducer was overlaid on the target. Placement of the transducer on the scalp was aided using a neuronavigation system (BrainSight, Rogue Research, Montreal, QUE, CAN).

#### Acoustic Modelling

Computational models were developed using individual subject MR and CT images to evaluate the wave propagation of LIFU across the skull and the resultant intracranial acoustic pressure maps. Simulations were performed using the k-Wave MATLAB toolbox(27), which uses a pseudospectral time domain method to solve discretized wave equations on a spatial grid. CT images were used to construct the acoustic model of the skull, while MR images were used to target LIFU at either the AI or aMCC target, based on individual brain anatomy. Details of the modeling parameters can be found in Legon et al. (2018)(28). CT and MR images were first co-registered and then resampled for acoustic simulations at a finer resolution and the acoustic parameters for simulation were calculated from the CT images. The skull was extracted manually using a threshold intensity value and the intracranial space was assumed to be homogenous as ultrasound reflections between soft tissues are small(29). Acoustic parameters were calculated from CT data assuming a linear relationship between skull porosity and the acoustic parameters(30,31). The computational model of the ultrasound transducer used in simulations was constructed to recreate empirical acoustic pressure maps of focused ultrasound transmitted in the acoustic test tank similar to previous work(29).

#### Model Heating

.Thermal models were performed using an adapted modified mixed-domain method (mSOUND)(32) with a 3-layer skull model (cortical-trabecular-cortical) from Benchmark 6 (33) to solve for the temperature fields based on the acoustic field and the bioheat equation using k-wave diffusion (27). This model assumes that 100% of the attenuation is absorption and thus converted to heat and, as such, is a conservative estimate of heating. We performed the thermal simulation for 30 ms on and 70 ms off (30% duty cycle) for 40 seconds using 1 MPa input pressure.

#### Sham condition

The Sham condition involved an active sham, where all procedures were identical to LIFU visits (including the use of neuronavigation, the selection of the proper gel puck, and the application of continuous auditory masking), except a high impedance material was inserted into the gel that attenuated the ultrasound prior to reaching the skull (34). For the Sham visit, the transducer was placed at one of the active sites, with the site randomized between participants. An auditory masking questionnaire was used after each session to query on successful masking.

#### Acoustic masking

In some cases, a single-element transducer can produce an audible auditory artifact likely as the result of the pulse repetition frequency. To address this potential confound (35), strict acoustic masking was performed. Acoustic masking was delivered through disposable earbuds that were plugged into a tablet. A combination of sounds from a white noise app were then randomly mixed creating a multitone that has previously been demonstrated to effectively mask ultrasound artifact (36). Participants were told to set the volume to a comfortable level that removed ambient sounds. This intensity range was on average 70 – 75 dB. Auditory masking was confirmed by speaking to the participant outside of their visual field. Auditory masking noise was played continuously throughout the testing session. 30 minutes after formal testing, participants were queried on auditory masking. Questions included “I could hear the LIFU stimulation”, “I could feel the LIFU stimulation”, and “I believe I experienced LIFU stimulation.” Participants were asked to respond to each question using a 7-point Likert scale (0–6) with points corresponding to Strongly Disagree / Disagree/ Somewhat Disagree / Neutral / Somewhat Agree / Agree / Strongly Agree.

### Physiology Preprocessing

#### Electromyography (EMG)

EMG recordings were bandpass filtered from 30 to 250 Hz using a 4^th^ order Butterworth filter and the Matlab function `filtfilt.` The data were then epoched around the startle events from -1 to 1 seconds. Each habituation block had 12 events. To quantify EMG magnitude within a defined post-stimulus interval, we used a custom MATLAB function. This function computed the rectified and baseline-corrected area under the curve (AUC) of a single-trial EMG trace from the time window [0.05 0.25] seconds. Each EMG waveform was first rectified to obtain its envelope and smoothed using a moving root-mean-square (RMS) window of 20 ms. The mean activity during a predefined baseline period [−0.2 0] seconds was then subtracted, and negative values were zero-floored to avoid spurious decreases. The resulting magnitude is integrated (trapezoidal numerical integration) over the time window [0.05 0.25] seconds, yielding a scalar measure of EMG response magnitude. The response magnitude from each event was used for quantification and statistical testing.

#### Electroencephalography (EEG)

EEG recordings were bandpass filtered from 0.1 to 55 Hz using a 3^rd^ order Butterworth filter and the Matlab function `filtfilt.` The data were then epoched around the startle events from -1 to 1 seconds. A similar approach to EMG was used to quantify EEG magnitude. We computed the rectified and baseline-corrected area under the curve (AUC) of a single-trial EEG trace from the time window [0.1 0.5] seconds. Each EEG waveform was first rectified to obtain its envelope and smoothed using a moving root-mean-square (RMS) window of 20 ms. The mean activity during a predefined baseline period [−0.2 0] seconds was then subtracted, and negative values were zero-floored to avoid spurious decreases. The resulting magnitude is integrated (trapezoidal numerical integration) over the time window [0.1 0.5] seconds, yielding a scalar measure of EEG response magnitude. The response magnitude from each event was used for quantification and statistical testing.

#### Electrodermal Response (EDR)

The electrodermal data stream was band-pass filtered from 0.5 to 5 Hz using a 2^nd^ order Butterworth filter and the Matlab function `filtfilt`. The data was then epoched around both the startle events and the aversive events from [-2 5] seconds. We computed the rectified and baseline-corrected area under the curve (AUC) of a single-trial EDR trace from the time window [0 5] seconds. Each EDR waveform was first rectified to obtain its envelope and smoothed using a moving root-mean-square (RMS) window of 20 ms. The mean activity during a predefined baseline period [−0.2 0] seconds was then subtracted, and negative values were zero-floored to avoid spurious decreases. The resulting magnitude is integrated (trapezoidal numerical integration) over the time window [0 5] seconds, yielding a scalar measure of EDR response magnitude. The response magnitude from each event was used for quantification and statistical testing.

### Analysis

We quantified EMG, EEG, and EDR startle habituation by computing trial-wise area-under-the-curve (AUC) responses. For each participant, LIFU session (AI, aMCC, Sham), and time point (Pre and Post LIFU), we estimated robust linear trends across the integer trial index [1:12] using the Theil–Sen median slope. Slopes were calculated separately for two unified windows: Early (trials 2–6) and Late (trials 7–12) with baseline AUC defined as the mean of Trial 1. The slope served as the dependent variable in all models, where more negative values indicate greater within-session attenuation of the startle response.

To control for baseline and anxiety covariates while separating within- from between-person effects, we decomposed both baseline AUC and state anxiety (STAI-S) using person-mean centering across the six LIFU × Time cells (AI, aMCC, Sham × Pre, Post). This yielded within-person deviations (Base_w, STAIS_w) and between-person means (Base_b, STAIS_b). Because STAI-S was measured once per session (i.e., no pre/post split), the session value was copied to both Pre and Post prior to centering. Trait anxiety (STAI-T) was grand-mean centered (STAI_Tc). We then tested LIFU effects on Early and Late slopes jointly using a linear mixed-effects model in MATLAB using `fitlme`, REML and Satterthwaite degrees of freedom for Type-III tests. The model was:

~~~
Slope ∼ LIFU * Time * Window + Base_w + Base_b + STAI_Tc + STAIS_w + STAIS_b + (1 + Time | Subj).
~~~

LIFU was coded with Sham as the reference (levels: Sham, AI, aMCC), Time included Pre and Post, and Window included Early and Late. Primary inference targeted the LIFU × Time × Window interaction, with planned Wald contrasts evaluating AI versus Sham and aMCC versus Sham for the Late Post–Pre change, the Early Post–Pre simple effects within AI and aMCC, and the “Late minus Early” change within each active condition. To visualize and model trial trajectories, we also fit a per-trial growth-curve mixed model with window-specific slopes and optional moderation by STAI-T, selecting the most complex estimable specification:

~~~
Y ∼ LIFU * Time + Window:Trial_c + (optional) LIFU:Window:Trial_c (± :STAI_Tc) + Base_w + Base_b + STAIS_w + STAIS_b + (1 + Trial_c | Subj).
~~~

Here, Trial_c is centered within window (Early or Late). Predicted trajectories were plotted at typical covariate values, and when the fitted model included STAI_Tc terms we displayed curves at the mean and at ±1 SD of STAI-T.

Before testing LIFU causation, we examined baseline associations between anxiety and habituation. For STAI-T (between-person), we collapsed Pre slopes across the three LIFU sessions to a per-subject mean for Early and Late and computed Spearman’s ρ with STAI-T; we also fit LMEs using all session-level Pre slopes

`(Early Slope ∼ STAI-T + (1 | Subj) and Late Slope ∼ STAI-T + (1 | Subj)`. For STAI-S (within-person), we estimated repeated-measures correlations between session-level STAI-S and Pre slopes (Early and Late) across the three sessions within each subject.

#### Cross-Modal Habituation Coupling Analysis

To assess whether habituation strength was coordinated across physiological modalities, we examined correlations among EMG, EEG, and EDR startle slopes derived from the analyses described above. For each modality, we used the Theil–Sen slope estimates for early (Trials 2–6) and late (Trials 7–12) habituation windows at pre- and post-LIFU timepoints. For each participant, the collapsed change in slope (Δ = Post – Pre) was extracted for both early and late windows, averaged across LIFU conditions, and aligned across modalities. Cross-subject coupling was quantified using Spearman’s rank correlations between EMG and EEG, and between EMG and EDR, computed separately for early and late windows. These analyses tested whether individuals showing greater habituation in one modality tended to show greater habituation in another.

To assess within-subject coupling across LIFU conditions, we computed session-specific Δ slopes (Post – Pre) for each subject and LIFU condition, aligned observations across modalities by subject and condition, and applied repeated-measures correlation to estimate within-person associations while accounting for between-subject variability.

We further evaluated whether cross-modal coupling was independent of anxiety by performing partial Spearman correlations between EMG and EEG Δ slopes and between EMG and EDR Δ slopes, controlling for both trait anxiety (STAI-T) and mean session-level state anxiety (STAI-S). In addition, confirmatory linear mixed-effects models were fit to quantify these relationships while adjusting for anxiety covariates, using the general form: `Outcome ∼ EMG + STAI-T + STAI-S + (1|Subj)`, where the outcome variable was EEG or EDR slope change. These models estimated the unique contribution of EMG slope changes to concurrent changes in cortical (EEG) or autonomic (EDR) habituation while accounting for within-subject dependencies and individual differences in anxiety.

Finally, to illustrate the overall association between modalities, we fit ordinary least-squares regression models to the collapsed Δ values across participants using EMG slope change as the predictor of EEG or EDR slope change (“total effect” analyses). These models provide an additional visualization of cross-modal coupling at the group level and were run separately for early and late habituation windows.

#### Mean Response Magnitude Analyses

As an additional line of inquiry (and more traditional analysis), we examined whether the mean magnitude of the first trial or trials 2:6 or 7:12 of the EMG, EEG, or EDR response correlated with either trait or state anxiety. Specifically, we tested whether individuals with higher trait anxiety (STAI-T) or higher session-specific state anxiety (STAI-S) showed stronger or weaker initial physiological responses (Trial 1) or if the mean magnitude across the Early or Late windows related to state or trait anxiety. We also assessed whether low-intensity focused ultrasound (LIFU) targeting the AI or aMCC, compared with Sham, altered the magnitude of this initial response or the mean response across each Early or Late window. This approach complemented our slope-based analyses by quantifying mean-level effects independent of trial-by-trial change.

To evaluate these relationships, for each physiological modality we extracted single-trial area-under-the-curve (AUC) magnitudes across the respective trials, 3 LIFU sessions (AI, aMCC, Sham), and 2 time points (pre, post) for each participant. State and trait anxiety measures were then merged so that there was one entry per Subject × LIFU × Time × Trial combination. Session-level STAI-S values were assigned to the corresponding LIFU condition for each participant, while the subject-level STAI-T value was constant across sessions.

For correlations, we first restricted analyses to the pre-LIFU baseline and computed (1) a between-person Spearman correlation between the average trial magnitude and STAI-T and (2) a within-person repeated-measures correlation between session-specific STAI-S and the corresponding mean physiological magnitude across LIFU conditions. The repeated measures correlation analysis was performed by centering both STAI-S and response magnitudes within each participant and estimating a common within-person slope, from which the correlation coefficient, t statistic, and p value were derived analytically. To assess mean-level treatment effects, we averaged trial magnitudes over user-defined trial windows (1 or 2:6 or 7:12) for each Subject × LIFU × Time combination and fit a linear mixed-effects model of the form:

~~~
Mean Magnitude ∼ 1 + LIFU * Time + (1∣Subj)
~~~

This model tested main and interaction effects of LIFU and Time while accounting for subject-level random intercepts. Post – Pre contrasts were computed for AI vs Sham and aMCC vs Sham, with corresponding t, p, and 95% confidence intervals. All analyses were implemented in MATLAB (MathWorks, Natick, MA) using custom scripts that automated data restructuring, correlation analyses (`corr` and `rmcorr`), mixed-effects model fitting (`fitlme`).

## RESULTS

### Ultrasound targeting

Group (N = 40) normalized and MNI registered ultrasound modelling showing distribution of pressure and overlap with the intended target is shown in **Figure 2**. Ultrasound beam characteristics, targeting depths, individual participant accuracy and auditory masking results are available in *Supplementary Information*.

**Figure 2.**
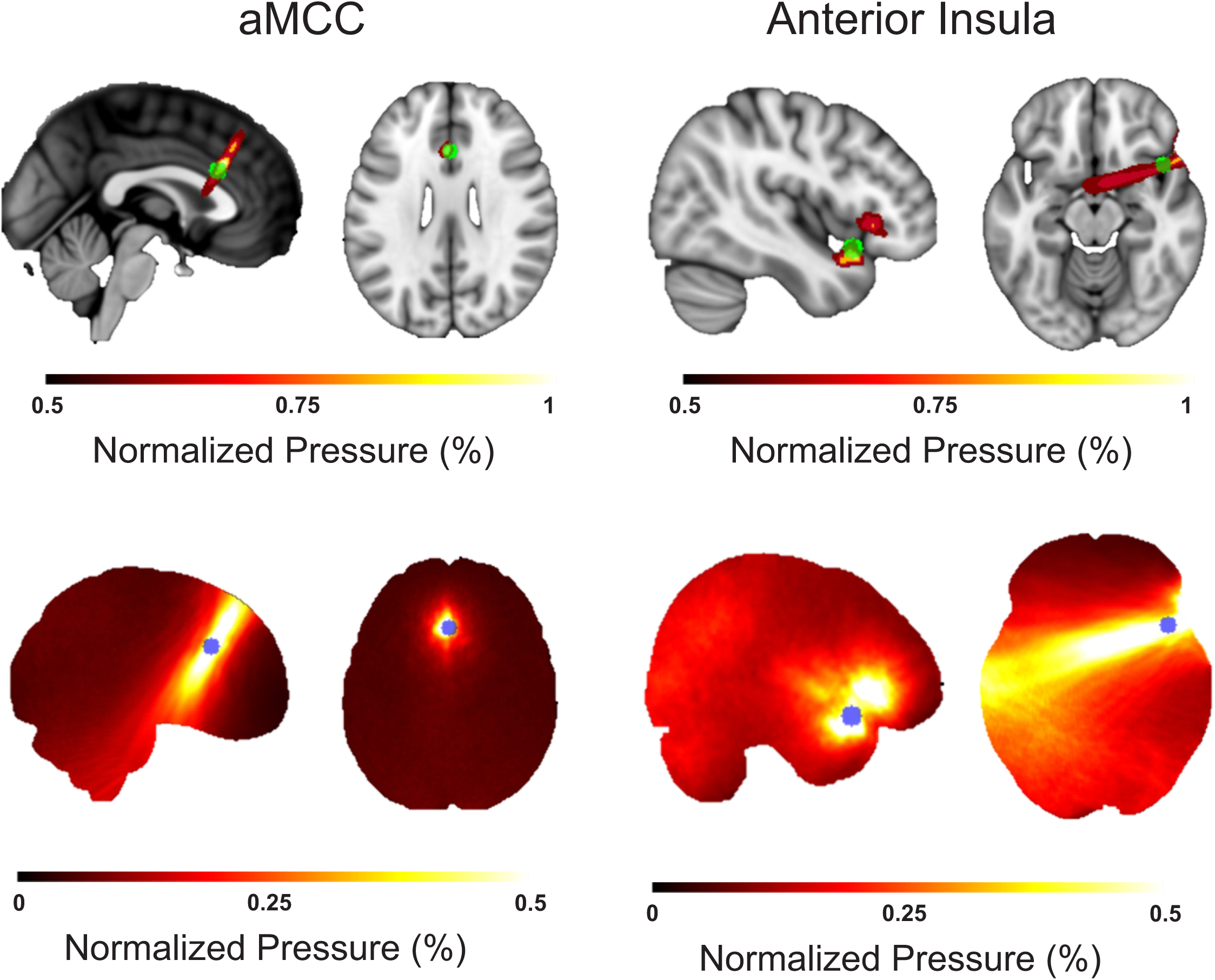
LIFU targeting the anterior insula and anterior mid-cingulate cortex (aMCC) (Left Top) Group (N = 40) normalized acoustic pressure (warm colors) showing top 50% pressure distribution in MNI space for anterior mid-cingulate cortex (aMCC). A 5mm ROI sphere (green) is shown centered on coordinate [0 22 26] to show targeting accuracy to scale. Scale bar is normalized pressure. (Left Bottome) Group (N = 40) normalized pressure map showing pressure distribution from 0 to 50% across the group at scale with above brain images. Scale bar is normalized pressure. (Right Top) Group (N = 40) normalized acoustic pressure (warm colors) showing top 50% energy distribution in MNI space for right anterior insula targeting to scale. A 5mm ROI sphere (green) is also shown centered on coordinate [44 14 -14] to show targeting accuracy to scale. Scale bar is normalized pressure. (Bottom Right) Group (N = 40) normalized pressure map showing pressure distribution from 0 to 50% across the group at scale with above brain images. Scale bar is normalized pressure.

### Trait Anxiety (STAI-T)

Participants filled out the STAI-T(37) on the first visit. The mean ± SD was 40.3 ± 11.3; the median was 39.5 with a range of 25 – 68 (see **Figure 3**).

**Figure 3.**
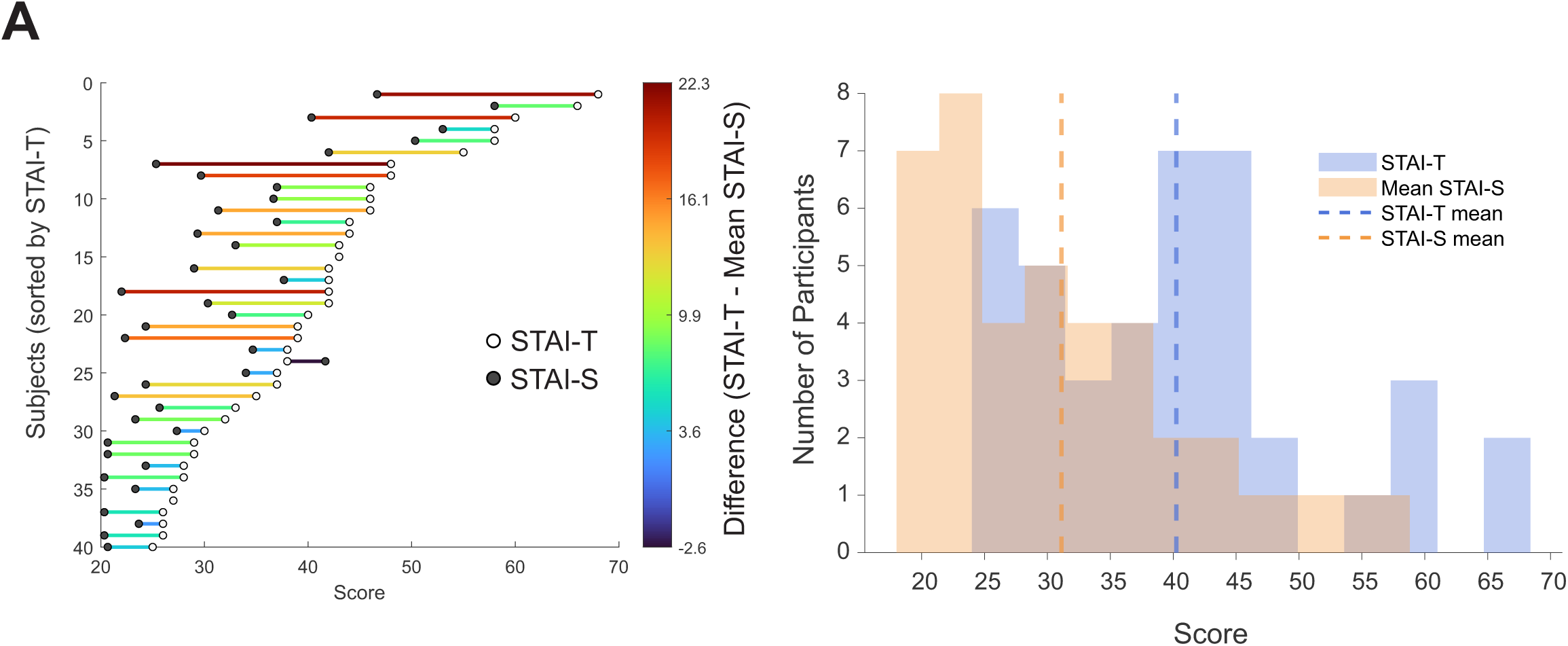
State and trait anxiety. **A.** (A) Individual dumbbell plots show each participant’s trait anxiety (STAI-T; right endpoint) and average state anxiety across sessions (mean STAI-S; left endpoint), sorted by descending STAI-T. Line color encodes each participant’s difference (STAI-T − mean STAI-S), with warmer hues indicating higher trait anxiety relative to their average state level. **B.** Histogram showing distribution of STAI-T scores in blue and mean STAI-S across sessions in orange. The mean of each is shown with the vertical dashed bars.

### State Anxiety (STAI-S)

Participants filled out the STAI-S 30 minutes before each formal LIFU session. The mean ± SD of all participants prior to LIFU for AI, aMCC and Sham was: 31.1 ± 11.4, 30.3 ± 9.8 and 31.9 ± 11.1. See **Figure 3** for barbell plot and histogram showing individual subject STAI-T and mean STAI-S across sessions.

#### EMG Startle Habituation and Relationship to Anxiety

Prior to LIFU, significant habituation was observed across trials (**Figure 4A & B**). The mean ± SEM AUC for Trial 1 collapsed across AI, aMCC and Sham condition before LIFU was: 32.0 ± 2.7; Trial 6: 15.9 ± 1.3 and Trial 12 was 12.5 ± 1.3. The mean ± SEM AUC for each trial for each condition pre/post LIFU are available in *Supplementary Information*.

**Figure 4.**
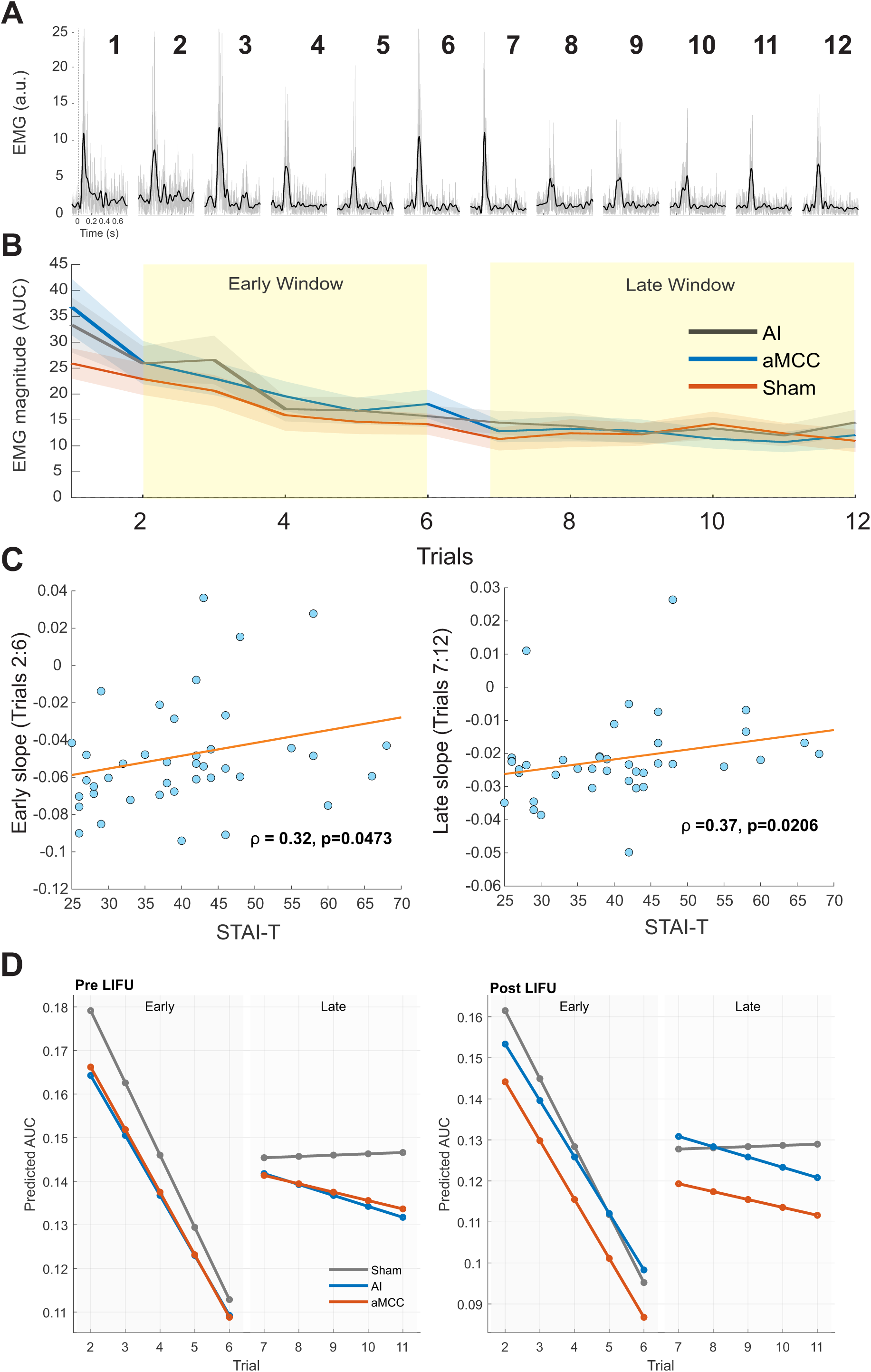
EMG startle results. **A.** Group (N = 40) rectified mean EMG responses across the twelve trials recorded from the orbicularis oculi muscle. The dark line is the mean and the grey is the raw data. Y-axis is EMG magnitude in arbitrary units (a.u). Time in seconds (s) is on the x-axis. The onset of the auditory startle is represented by the vertical dashed line at time 0. **B.** Group (N = 40) mean ± SEM EMG AUC plotted over trials prior to LIFU intervention. Trial 1 was used as baseline; Trials 2-6 were designated as the Early phase and Trials 7-12 were designated the Late phase. **C.** Pre-LIFU habituation slopes versus anxiety (Spearman). For each phase (Early: Trials 2–6; Late: Trials 7–12), the two panels summarize rank-based associations between startle habituation and STAI-T anxiety scores. Left: STAI-T vs. pre-session slope (Theil–Sen) collapsed across the three LIFU sessions (AI, aMCC, Sham) by subject. Points are subjects, the overlaid line is an OLS trend for visualization. Spearman’s ρ and two-sided p are provided. Slopes are computed with the Theil–Sen estimator from trial-wise AUCs; no baseline or post-session data enter these plots**. D.** Model-predicted trial trajectories by LIFU and session (lines-only). Left: Pre; Right: Post. For each LIFU condition (Sham, gray; AI, blue; aMCC, orange), solid lines show the linear within-phase trends predicted by the growth-curve linear mixed-effects model across the Early phase (Trials 2–6) and Late phase (Trials 7–12). Predictions were generated from the fitted model with phase-specific trial slopes and covariates held at typical values (within-subject Baseline and STAI-S = 0; between-subject Baseline and STAI-S = their sample means. Markers represent per-trial predicted AUC. Slope for Early vs. Late phases significantly differed (p < 0.05) but LIFU did not affect slopes for either phase.

Before LIFU, individual differences in trait anxiety were related to startle habituation. Across participants, higher STAI-T scores were associated with less negative (i.e., weaker) slopes during both early and late windows of the habituation block (Spearman’s ρ = 0.32, p = 0.047 for early; ρ = 0.37, p = 0.021 for late) (**Figure 4C**). Linear mixed-effects models including random subject intercepts produced similar, albeit nonsignificant, trends for trait anxiety (early: β = 0.00069, p = 0.086; late: β = 0.00030, p = 0.114). In contrast, within-subject fluctuations in state anxiety (STAI-S) were not significantly related to early (rmcorr r = 0.15, p = 0.19) or late (rmcorr r = 0.04, p = 0.70) habituation slopes.

The primary mixed-effects model tested the effects of LIFU condition (AI, aMCC, Sham), Time (Pre, Post), and Window (Early, Late), controlling for baseline startle magnitude (Base_w, Base_b), state anxiety (STAIS_w, STAIS_b), and trait anxiety (STAI_Tc), with subject random intercepts. The overall model revealed a robust main effect of Window (F(1,463) = 13.52, p < 0.001), indicating that late-phase slopes were significantly more positive (i.e., less habituated) than early slopes across all conditions. Baseline covariates were also significant, with both between-subject mean baseline amplitude (F(1,463) = 17.97, p < 0.001) and within-session deviations (F(1,463) = 3.07, p = 0.08) predicting slope magnitude, suggesting that participants with larger baseline startle responses showed reduced habituation. Between-person state anxiety (STAIS_b) was modestly associated with slope: F(1,463) = 5.25, p = 0.022, whereas neither within-person STAI-S nor trait anxiety (STAI_Tc) significantly contributed to the model.

No significant main or interaction effects involving LIFU were observed. The LIFU × Time × Window interaction was nonsignificant (F(2,463) = 0.16, p = 0.85), and planned contrasts confirmed that changes in habituation from Pre to Post did not differ between AI or aMCC stimulation and Sham in either early or late phases (all ps > 0.24). Likewise, the “Late minus Early” change within AI and aMCC did not differ from Sham (all ps > 0.57). Thus, EMG startle habituation slopes were stable across sessions and unaffected by LIFU.

The growth-curve mixed model incorporating trial-wise data successfully converged using the full specification: `Y ∼ LIFU*Time + Window:Trial_c + LIFU:Window:Trial_c + LIFU:Window:Trial_c:STAI_Tc + Base_w + Base_b + STAIS_w + STAIS_b + (1 + Trial_c|Subj))`. This model reproduced the main finding that habituation was steeper during early than late trials, with no systematic modulation by LIFU condition, Time, or their interactions with trait anxiety (**Figure 4D**). Overall, EMG startle responses exhibited clear habituation within windows, with shallower slopes in later trials and modest associations with trait but not state anxiety. LIFU to the anterior insula or anterior mid-cingulate cortex did not alter habituation magnitude relative to sham.

#### EEG Startle Habituation and Relationship to Anxiety

Prior to LIFU, significant habituation was observed across trials (**Figure 5A & B**). The mean ± SEM for Trial 1 collapsed across AI, aMCC and Sham condition before LIFU was: 34.3 ± 4.7; Trial 6: 14.7 ± 0.9 and Trial 12 was 15.0 ± 1.1. The mean ± SEM for each trial for each condition pre/post LIFU are available in *Supplementary Information*. Before LIFU, no significant relationships emerged between anxiety and EEG habituation slopes. Across participants, trait anxiety (STAI-T) was unrelated to either early (ρ = –0.08, p = 0.62) or late (ρ = 0.19, p = 0.24) slopes when collapsed across sessions. Linear mixed-effects models that included random subject intercepts likewise showed no significant associations between STAI-T and early (β = – 0.00042, p = 0.57) or late (β = 0.00016, p = 0.43) habituation slopes. Within-subject fluctuations in state anxiety (STAI-S) were also unrelated to EEG slopes at either window (rmcorr r = –0.03, p = 0.78 for early; r = 0.03, p = 0.81 for late), indicating no consistent influence of state anxiety on cortical habituation prior to LIFU. The full mixed-effects model tested LIFU condition (AI, aMCC, Sham), Time (Pre, Post), and Window (Early, Late) while controlling for baseline EEG amplitude (Base_w, Base_b), state anxiety (STAIS_w, STAIS_b), and trait anxiety (STAI_Tc), with random subject intercepts. The model revealed a robust main effect of Window: F(1,463) = 20.26, p < 0.001, showing that late-window EEG slopes were significantly more positive than early-window slopes, consistent with reduced cortical habituation across trials. There was also a significant main effect of Time: F(1,463) = 6.67, p = 0.010; with overall slopes becoming more positive at Post compared to Pre, suggesting weaker habituation following LIFU sessions irrespective of condition. Baseline AUC strongly predicted slope magnitude (Base_w: F(1,463) = 24.04, p < 0.001; Base_b: F(1,463) = 31.44, p < 0.001), indicating that individuals with larger baseline responses exhibited smaller trial-wise decreases. No significant effects were observed for within- or between-person state anxiety or for trait anxiety. Although no overall LIFU × Time × Window interaction reached significance: F(2,463) = 2.03, p = 0.13, a LIFU × Window effect was detected: F(2,463) = 3.20, p = 0.042, reflecting subtle baseline differences in the pattern of early versus late EEG habituation across LIFU conditions. Planned contrasts confirmed that neither AI nor aMCC stimulation altered habituation relative to Sham for either early or late windows (all ps > 0.12). However, there was a trend toward a greater Late-versus-Early change in AI: F(1,463) = 3.45, p = 0.064, suggesting a possible but nonsignificant enhancement of late-window attenuation (**Figure 5C**). The per-trial growth-curve model successfully converged with the full specification. This model replicated the main findings of steeper early-window than late-window habituation and a general post-session reduction in slope, without evidence for significant modulation by LIFU or anxiety covariates (**Figure 5C**). Overall, EEG startle responses demonstrated robust within-block habituation that was stronger early in the block and diminished across trials. Habituation was modestly reduced after LIFU sessions but did not differ significantly between active and sham conditions, nor was it systematically related to trait or state anxiety.

**Figure 5.**
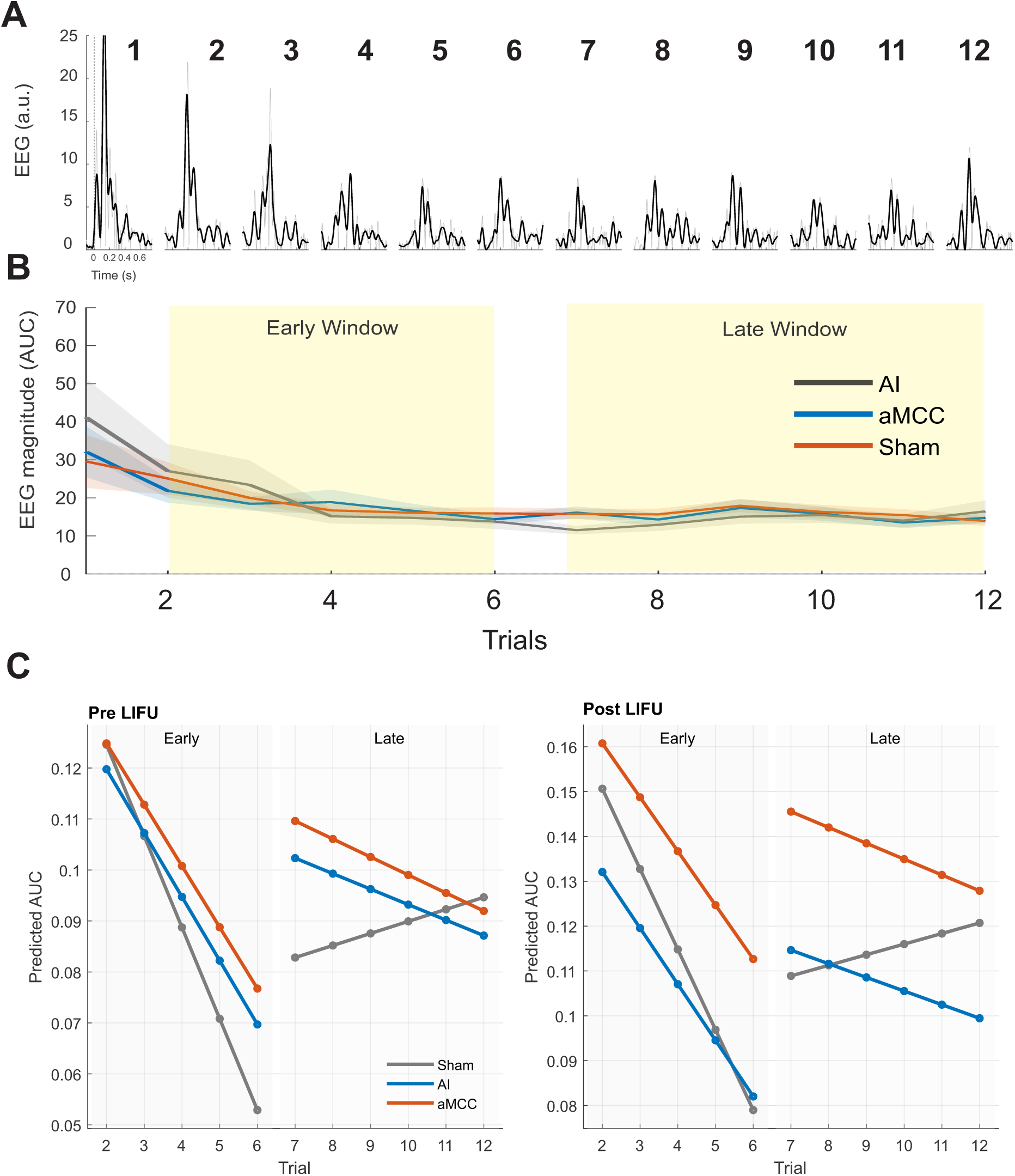
EEG startle results. **A.** Group (N = 40) rectified mean EEG responses across the twelve trials recorded from scalp electrode CZ. The dark line is the mean and the grey is the raw data. Y-axis is EEG in arbitrary units (a.u). Time in seconds (s) is on the x-axis. The onset of the auditory startle is represented by the vertical dashed line at time 0. **B.** Group (N = 40) mean ± SEM EEG AUC plotted over trials prior to LIFU intervention. Trial 1 was used as baseline; Trials 2-6 were designated as the Early window and Trials 7-12 were designated the Late window. **C.** Model-predicted trial trajectories by LIFU and session (lines-only). Left: Pre; Right: Post. For each LIFU condition (Sham, gray; AI, blue; aMCC, orange), solid lines show the linear within-window trends predicted by the growth-curve linear mixed-effects model across the Early (Trials 2–6) and Late (Trials 7–12) windows. Predictions were generated from the fitted model with window-specific trial slopes and covariates held at typical values (within-subject Baseline and STAI-S = 0; between-subject Baseline and STAI-S = their sample means; STAI-T centered at its mean when included). Markers represent per-trial predicted AUC. LIFU did not affect slopes in either the early or late window and EEG slopes in either early or late windows were not associated with state or trait anxiety scores despite demonstrate similar slope dynamics to the EMG data.

#### EDR Startle Habituation and Relationship to Anxiety

Prior to LIFU, significant habituation was observed across trials (**Figure 6A**). The mean ± SEM for Trial 1 collapsed across AI, aMCC and Sham condition before LIFU was: 69.5 ± 9.6; Trial 6: 13.7 ± 2.5 and Trial 12 was 7.6 ± 2.1. The mean ± SEM for each trial for each condition pre/post LIFU are available in *Supplementary Information*.

**Figure 6.**
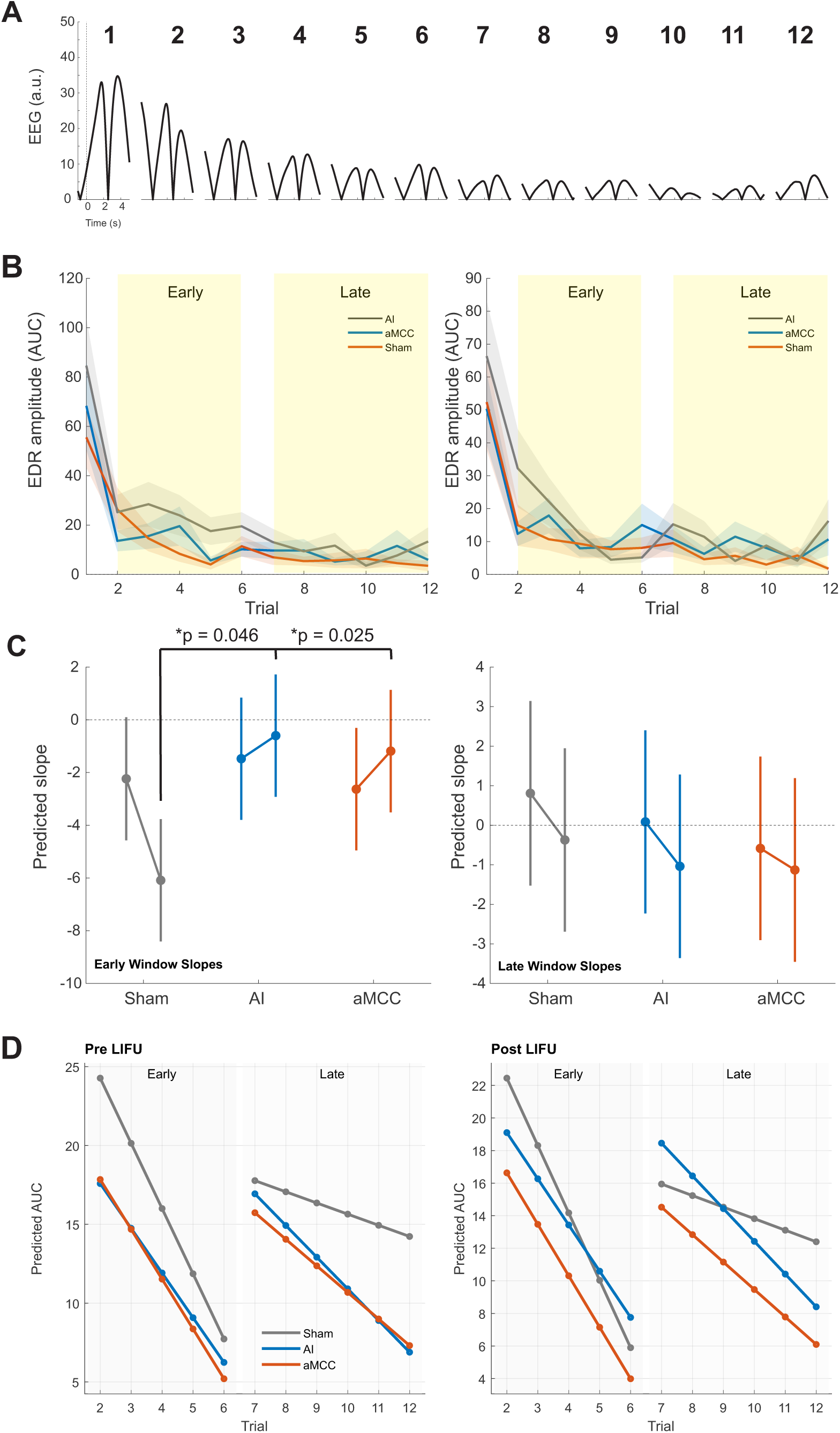
EDR startle results. **A.** Group (N = 40) rectified mean EDR responses across the twelve trials recorded from the hand. The dark line is the mean and the grey is the raw data. Y-axis is EEG in arbitrary units (a.u). Time in seconds (s) is on the x-axis. The onset of the auditory startle is represented by the vertical dashed line at time 0. **B.** Group (N = 40) Trial-wise EDR amplitudes by LIFU condition and session. Mean (± SEM) EDR amplitudes are plotted across 12 habituation trials for each LIFU condition (AI, blue; aMCC, orange; Sham, gray) during the Pre (left) and Post (right) sessions. Shaded bands represent ±1 SEM across subjects. Light gray vertical regions indicate the Early (Trials 2–6) and Late (Trials 7–12) analysis windows used in slope estimation. Responses are expressed as trial-wise AUC values averaged across subjects. **C.** Model-based predicted slopes (95% CI) by LIFU condition and session. Predicted mean Early and Late slopes from the mixed-effects model are shown for each LIFU condition (Sham, gray; AI, blue; aMCC, orange) across Pre and Post sessions. Bars depict model-estimated means ± 95% CIs, holding within-subject covariates at 0 and between-subject covariates at their sample means; STAI-T was centered at the mean. The model revealed a significant LIFU × Time interaction: F(2,463)=3.04, p=0.049. Planned contrasts showed that both AI and aMCC stimulation increased Early-window slope change relative to Sham (AI vs Sham: F(1,463)=4.02, p=0.046; aMCC vs Sham: F(1,463)=5.04, p=0.025), whereas Late-window changes were not significant (all p > 0.78). Baseline covariates contributed strongly (Within: F=12.29, p=0.0005; Between: F=26.94, p<0.001), whereas state and trait anxiety were non-significant (all p > 0.56). **D.** Model-predicted trial trajectories by LIFU and session (lines-only). Left: Pre; Right: Post. For each LIFU condition (Sham, gray; AI, blue; aMCC, orange), solid lines show the linear within-window trends predicted by the growth-curve linear mixed-effects model across the Early (Trials 2–6) and Late (Trials 7–12) windows. Predictions were generated from the fitted model with window-specific trial slopes and covariates held at typical values (within-subject Baseline and STAI-S = 0; between-subject Baseline and STAI-S = their sample means; STAI-T centered at its mean when included). Markers represent per-trial predicted AUC. LIFU to AI and aMCC increased the early-window slope compared to Sham. Together, these results indicate that LIFU to AI and aMCC selectively enhanced Early-phase habituation slopes compared to Sham, consistent with greater initial modulation of physiological reactivity following active stimulation.

Before LIFU, electrodermal responses showed no reliable association with either trait or state anxiety. Across participants, correlations between STAI-T and early (ρ = –0.09, p = 0.60) or late (ρ = –0.21, p = 0.21) slopes were nonsignificant, and linear mixed-effects models including random subject intercepts confirmed that trait anxiety did not predict EDR habituation (early: β = 0.10, p = 0.21; late: β = –0.03, p = 0.28). Within-subject fluctuations in state anxiety (STAI-S) were similarly unrelated to either early (rmcorr r = 0.07, p = 0.56) or late (rmcorr r = 0.01, p = 0.95) slopes, indicating that autonomic habituation before stimulation was not influenced by anxiety levels.

The primary mixed-effects model tested the effects of LIFU condition (AI, aMCC, Sham), Time (Pre, Post), and Window (Early, Late) on EDR slopes, controlling for baseline response magnitude (Base_w, Base_b), state anxiety (STAIS_w, STAIS_b), and trait anxiety (STAI_Tc), with random intercepts for subjects. The model revealed a significant main effect of Time: F(1,463) = 5.32, p = 0.022, indicating overall differences in habituation slopes from Pre to Post, and a marginal effect of Window: F(1,463) = 3.34, p = 0.068, suggesting slightly steeper early than late slopes. Baseline electrodermal activity was a strong predictor of slope (Base_w: F(1,463) = 12.29, p < 0.001; Base_b: F(1,463) = 26.94, p < 0.001), consistent with reduced habituation in individuals showing larger baseline amplitudes. Neither STAI-S nor STAI-T significantly predicted slope magnitude. Importantly, a LIFU × Time interaction reached significance: F(2,463) = 3.04, p = 0.049, suggesting differential pre–post changes across stimulation sites. Planned contrasts indicated that early-window habituation slopes became more negative (i.e., stronger habituation) following LIFU to AI: F(1,463) = 4.02, p = 0.046 and aMCC: F(1,463) = 5.04, p = 0.025 relative to sham (**Figure 6B & C**), while no significant effects were observed for the late-window or for the “Late minus Early” difference (all ps > 0.16). Thus, active LIFU to either target enhanced early-window autonomic habituation but did not affect later trials. The per-trial growth-curve model converged successfully and reproduced these findings: steeper early-window habituation and enhanced early attenuation after active LIFU, without evidence of modulation by anxiety covariates **(Figure 6D**). Overall, EDR startle responses showed modest within-block habituation that was strongest early in the block and significantly increased following AI and aMCC stimulation compared to sham. These results suggest that LIFU may transiently augment early-window autonomic adaptation to repeated startle probes, independent of baseline anxiety.

#### Cross-Modal Habituation Coupling

To determine whether habituation strength was coordinated across physiological systems, we examined cross-modal correlations among EMG, EEG, and EDR slope changes (Δ = Post – Pre) for both early and late habituation windows. Across subjects, EMG and EEG slope changes were strongly positively correlated for both late (ρ = 0.42, p = 0.0076) and early (ρ = 0.42, p = 0.0083) windows (**Figure 7A & B**), indicating that individuals who exhibited greater startle habituation in EMG also showed greater cortical habituation in EEG. In contrast, EMG and EDR slopes were not significantly related in either window (late: ρ = 0.02, p = 0.89; early: ρ = 0.21, p = 0.19), suggesting that autonomic habituation did not systematically track somatic or cortical measures of startle attenuation across participants (**Figure 7A & B**).

**Figure 7.**
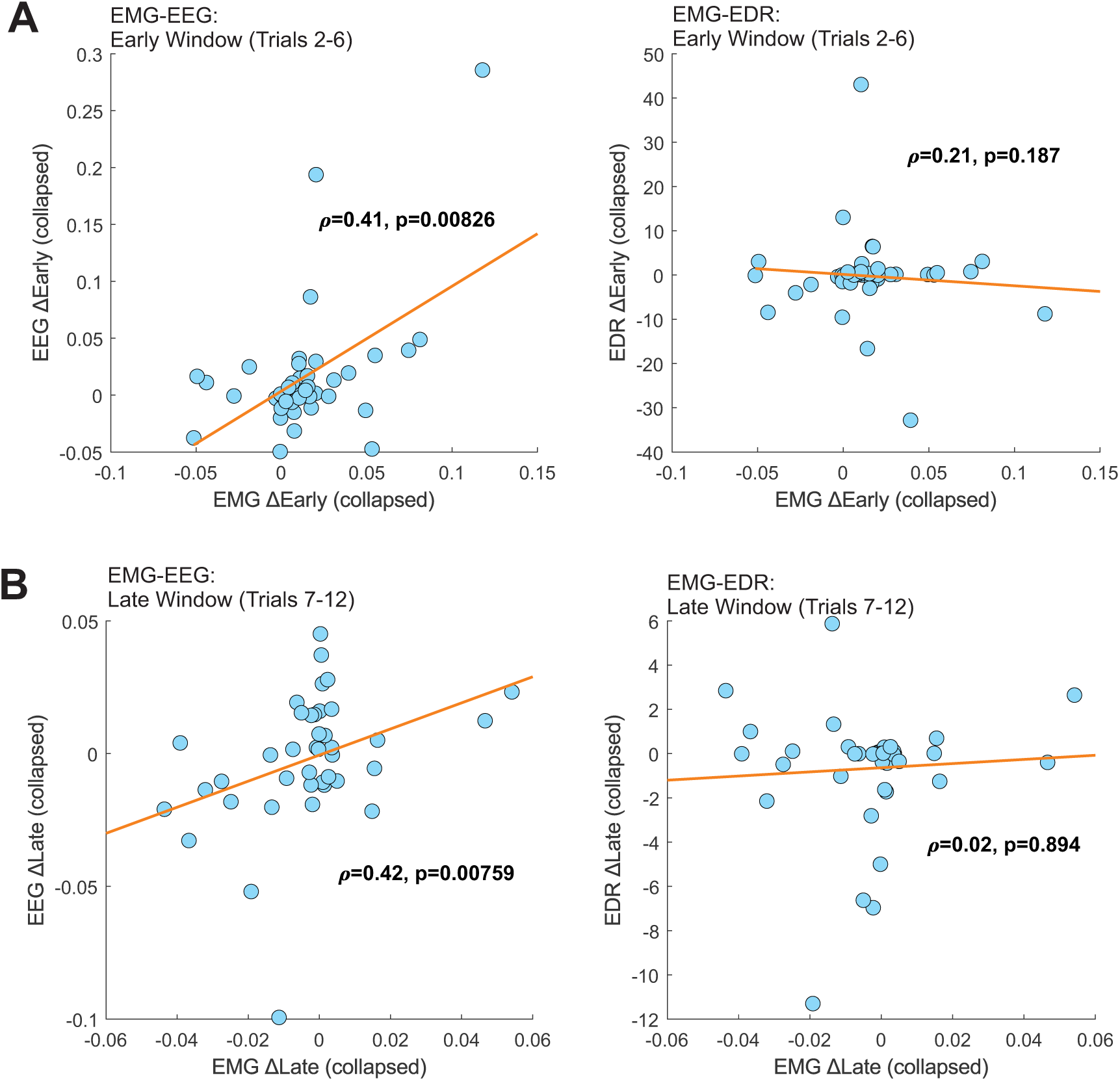
Cross-modal coupling of habituation slopes. **A.** *Early slopes*. Panels show cross-subject associations in change (Post–Pre) of habituation slope across modalities, collapsed across LIFU conditions. Each point represents one participant’s ΔEarly slope, computed from Theil–Sen fits to trial-wise startle or physiological responses. Left: EMG–EEG coupling; Right: EMG–EDR coupling. Lines indicate least-squares fits for visualization. Values denote Spearman’s ρ and p-values. A positive ρ indicates that individuals who showed greater post-session slope change in one modality also tended to show greater change in the other. The Early window corresponds to Trials 2–6. **B.** *Late slopes*. Panels show cross-subject associations in change (Post–Pre) of habituation slope across modalities, collapsed across LIFU conditions. Each point represents one participant’s ΔLate slope, computed from Theil–Sen fits to trial-wise startle or physiological responses. Left: EMG–EEG coupling; Right: EMG–EDR coupling. Lines indicate least-squares fits for visualization. Values denote Spearman’s ρ and p-values. A positive ρ indicates that individuals who showed greater post-session slope change in one modality also tended to show greater change in the other. The Late window corresponds to Trials 7–12.

Repeated-measures correlations examining within-subject coupling across LIFU sessions were indeterminate due to insufficient within-person variability. However, when anxiety covariates were included, the partial Spearman correlation between EMG and EEG remained robust (late: ρ = 0.46, p = 0.0037; early: ρ = 0.39, p = 0.016), demonstrating that the cross-modal association was independent of both trait (STAI-T) and state (STAI-S) anxiety. Consistent results were observed in the linear mixed-effects models controlling for anxiety (EEG ∼ EMG + STAI-T + STAI-S + (1|Subj)). EMG slopes significantly predicted EEG slopes (late: β = 0.53, p = 0.015; early: β = 0.92, p = 0.0013), whereas neither trait nor state anxiety contributed to the model. These models accounted for subject-level random effects and confirmed that greater EMG habituation was associated with stronger EEG habituation.

Finally, simple regression models of the total effect corroborated these findings: EMG slopes significantly predicted EEG slopes across subjects for both late (β = 0.49, p = 0.018, R² = 0.14) and early (β = 0.92, p < 0.001, R² = 0.26) windows. No comparable associations emerged for EMG–EDR pairs.

Overall, cross-modal analyses revealed a consistent and anxiety-independent coupling between EMG and EEG habituation strength, suggesting that cortical and somatic indices of startle attenuation reflect a shared neural adaptation process. In contrast, electrodermal (EDR) habituation varied independently, implying that autonomic adaptation to repeated startle stimuli is partially dissociable from motor and cortical habituation mechanisms.

### Mean Response Magnitude Analyses

None of EMG, EEG or EDR Trial 1 or trials 2:6 or trials 7:12 magnitude significantly correlated with STAI-S or STAI-T scores (all p-corrected ≥ 0.05) (see Supplemental Table 1). None of the EMG, EEG or EDR Trial 1 or trials 2:6 or 7:12 mean magnitudes were affected by LIFU to either the AI vs. Sham (all p ≥ 0.10) or aMCC vs. Sham (all p ≥ 0.43). See Supplemental Table 1.

## Discussion

The present study examined whether anxiety is associated with deficits in sensory inhibition as indexed by the habituation slope of the acoustic startle response and whether transient modulation of salience-network hubs (AI or aMCC) via low-intensity focused ultrasound (LIFU) could alter these dynamics. Using simultaneous somatic (EMG), cortical (EEG), and autonomic (EDR) recordings across randomized AI, aMCC, and Sham sessions, we quantified the rate of habituation across trials, rather than relying on mean response magnitude. Habituation was modeled using robust Theil–Sen slopes for early (Trials 2–6) and late (Trials 7–12) windows and tested with mixed-effects models that parsed within- and between-person variance in baseline activity and anxiety (STAI-S, STAI-T). Overall, the data partially supported our initial hypotheses. First, consistent with our prediction that anxiety would relate to reduced sensory inhibition, we observed that higher trait anxiety was modestly associated with weaker EMG habituation across both early and late windows (ρ ≈ 0.3), indicating slower somatic adaptation in individuals with higher trait anxiety. However, this relationship did not extend to EEG or EDR measures, and state anxiety showed no reliable associations with habituation in any modality. Thus, the trait anxiety–inhibition link was supported for the somatic (EMG) channel only, suggesting that anxiety-related impairment in habituation to repeated provoking stimuli may manifest in reflex circuitry rather than in cortical or autonomic domains. The EMG response to startle stimuli is mediated by a conserved reflex circuit centered on the caudal pontine reticular nucleus (PnC), the core relay of the blink pathway (auditory nerve → cochlear nucleus → PnC → spinal interneurons → facial motor neurons) that receives extensive modulatory input from limbic and prefrontal regions (38,39). Affective states influence this brainstem arc through projections from the extended amygdala and associated forebrain structures. The central amygdala mediates phasic, cue-bound potentiation of startle, whereas the bed nucleus of the stria terminalis (BNST) sustains responses to uncertain threat—an anxiety-relevant mode that also modulates reflex output (14). Neuroimaging and lesion data show coactivation of the amygdala and anterior cingulate cortex during affective startle tasks, implicating top-down cortical regulation of this subcortical circuit (40). Recent simultaneous EMG–fMRI work demonstrates trial-by-trial coupling between PnC and centromedial amygdala activation, confirming a conserved amygdala–PnC pathway for defensive responding (41). Moreover, engagement of the anterior insula, anterior/mid-cingulate, orbitofrontal cortex, and cerebellum during startle modulation links this reflex system to salience and emotion-regulation networks implicated in anxiety (40,41). Within this integrated framework, our observation that higher trait anxiety predicts weaker EMG habituation suggests that reduced cortical inhibition or heightened amygdala–BNST drive may slow somatic adaptation to repeated aversive probes, consistent with models distinguishing phasic “fear” (amygdala-centric) from sustained “anxiety” (BNST-centric) processes (14,42). Interestingly, mean response magnitudes as quantified from the first trial or the mean of early or late window trials were unrelated to anxiety reinforcing that habituation slopes, not overall amplitudes, better capture anxiety-relevant inhibitory mechanisms.

Second, we hypothesized that LIFU to the anterior insula or anterior mid-cingulate would normalize or enhance habituation, particularly for cortical (EEG) and autonomic (EDR) responses based upon 1) both AI and aMCC being primary hubs of both the salience network (43,44) and central autonomic networks (45,46) and 2) that our prior work has demonstrated LIFU to the AI and aMCC reduce the amplitude of aversive nociceptive evoked responses (47,48). This hypothesis was partially confirmed: LIFU produced a selective, window-specific effect on autonomic adaptation only. Both AI and aMCC modulation significantly increased early-window EDR habituation relative to sham, yielding steeper (more negative) early slopes, while leaving EMG and EEG slopes unchanged. These findings suggest that LIFU transiently facilitates autonomic habituation during the most plastic early window of adaptation, potentially through engagement of salience-network control of visceromotor circuits. Electrodermal responses are driven by sympathetic efferents originating in hypothalamic and brainstem nuclei but are strongly modulated by higher cortical and limbic regions that integrate emotion, attention, and bodily arousal (49). Functional imaging and lesion studies identify a distributed “central autonomic network” comprising the anterior insula, anterior and mid-cingulate cortices, ventromedial prefrontal cortex, amygdala, and parietal regions that govern and represent EDR fluctuations(49). Within this hierarchy, the anterior insula provides viscerosensory representation of internal arousal states, while the aMCC integrates these bodily signals with cognitive and motivational context to adapt behavioral responses(49). LIFU-induced enhancement of early-window EDR habituation may reflect modulation of this salience–autonomic interface, perhaps through transient inhibition that increases cortical control over sympathetic output to a provoking stimulus. The lack of LIFU effect on the EEG response is curious and contradictory to our original hypotheses. The surface EEG response to a brief provoking stimulus is a multi-modal brain response that reflects the salience of the stimulus (50) as well as defensive response to it (51,52). Both the anterior insula and aMCC contribute to this potential and thus we hypothesized that LIFU would increase habituation slopes as a marker of more rapid habituation dynamics. Our previous work demonstrated LIFU effects on mean vertex responses to aversive nociceptive stimuli but perhaps the lack of effect here is due to a lack of required behavioral response (i.e. intensity or aversive reporting) such that the stimulus itself lack behavioral relevance and thus the cortical EEG marker of this was left unaffected. Nevertheless, the absence of LIFU effects on EEG or EMG modulation indicates that LIFU effects were not global and did not generalize to cortical or somatic habituation as initially hypothesized.

Third, although not a primary hypothesis, cross-modal analyses revealed a robust and anxiety-independent coupling between EMG and EEG habituation (ρ ≈ 0.4–0.5), suggesting coordinated cortical–somatic adaptation processes to repetitive provoking stimuli. In contrast, EDR habituation was largely uncorrelated with EMG, underscoring partial independence of autonomic adaptation mechanisms. This dissociation reinforces the interpretation that the observed LIFU effects on EDR represent targeted modulation of a distinct, autonomic component of sensory inhibition rather than a broad system-wide change.

Taken together, these results demonstrate that anxiety influences the rate of habituation, not the magnitude of startle responses, and primarily through somatic pathways. LIFU to salience-network hubs selectively enhanced early-window autonomic habituation, offering partial causal support for the proposed role of these regions in brain and body responses to provoking stimuli. Thus, the data align with the overall theoretical framework but reveal modality-specific and window-restricted effects highlighting EMG habituation as an anxiety-linked biomarker and EDR habituation as a potential LIFU-sensitive index of autonomic adaptation. Collectively, the findings emphasize that defensive sensory inhibition is a multi-component process, only some of which are altered by anxiety or modifiable through noninvasive neuromodulation.

## Data Availability

All data produced in the present study are available upon reasonable request to the authors

## Acknowledgements

This study would like to thank Jessica Florig and Kathryn Painchaud for help with data collection. This work was funded by a grant to WL from the Focused Ultrasound Foundation.

## Supplementary Information

### Ultrasound beam characteristics

The ultrasound beam as measured in free water had a lateral full-width at half maximum (FWHM) resolution in the X plane of 3.3 mm and in the Y plane of 3.4 mm at the Z maximum. FWHM represents the area that received > 50% of peak energy. The axial FWHM was 23 mm ranging from - 10 mm to + 13 mm from the point of maximum pressure (38 mm from the exit plane) conferring an effective axial FWHM of 28 – 51 mm. The constructed model waveform used for all acoustic simulations was in good agreement with these empirical measurements validating its use in the models as has been demonstrated previously for both insula and aMCC targets (36–38).

### Ultrasound heating

The maximum heating was 37.8 ℃ in trabecular bone (Δ 0.8 ℃) and 37.4 ℃ (Δ 0.4 ℃) in the brain.

### Targeting Depths & Accuracy

.The average depth of the AI target for males and females was: 39.0 ± 2.8 mm and 36.4 ± 3.4 mm respectively. The average depth of the aMCC target for males and females was: 49.9 ± 3.2 mm and 46.7.4 ± 2.5 mm.

The mean ± SD of the transducer placement on the head from the prescribed spot for right AI and aMCC was: 0.64 ± 0.63 mm and 0.87 ± 0.73 mm respectively. A paired t-test revealed no significant differences t(39) = 1.51, p = 0.14. (See **Supplementary Figures 1 & 2**).

**Supplementary Figure 1.**
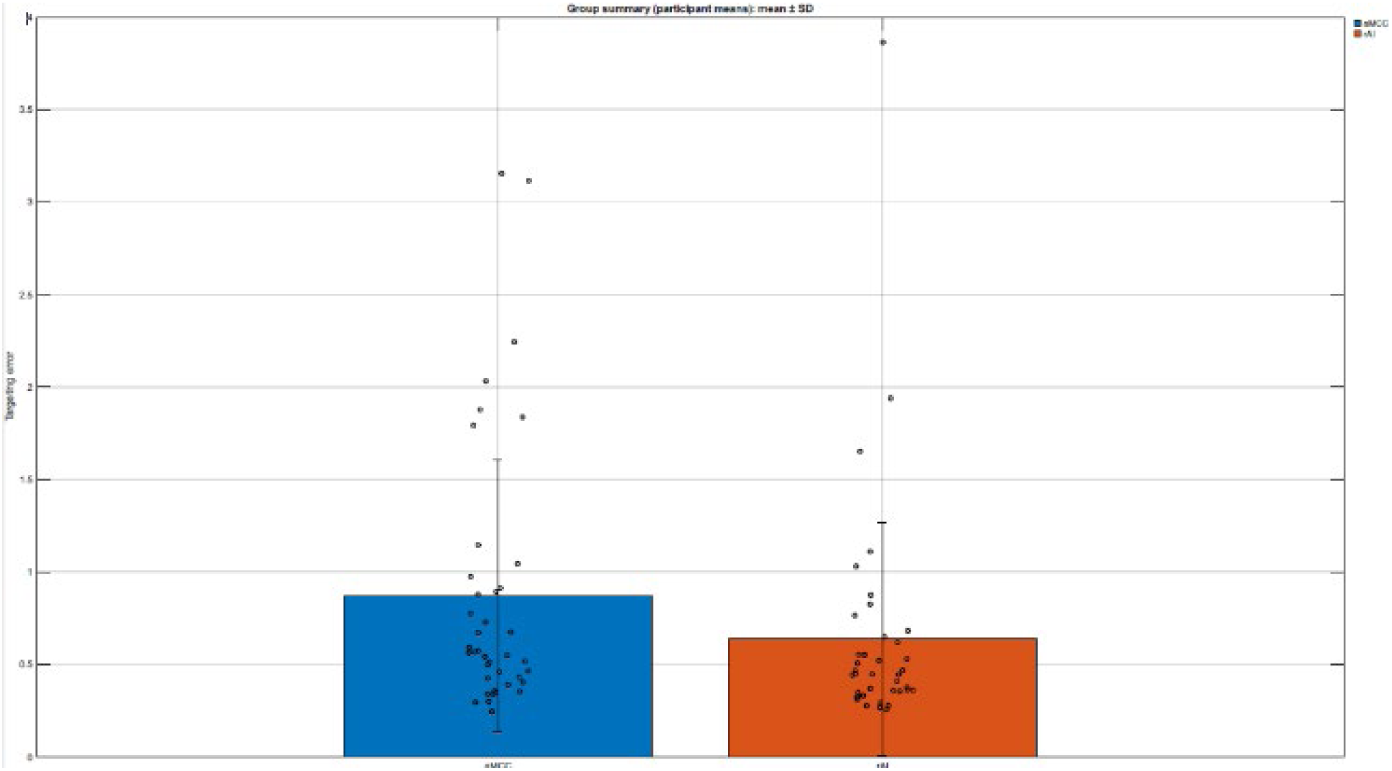

**Supplementary Figure 2.**
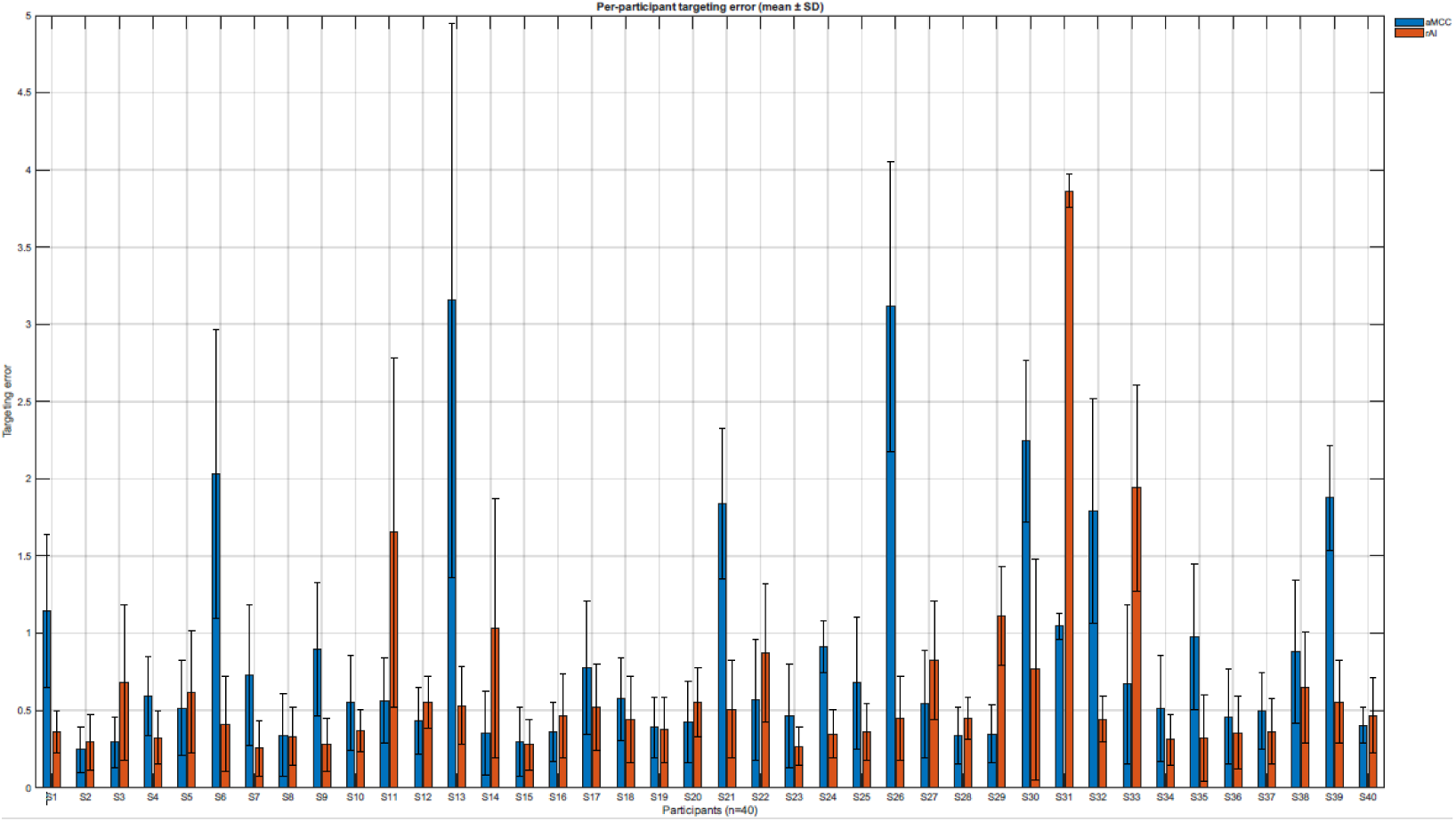

### Auditory Masking

We converted participant responses into numerical values 0 – 6 where 0 = strongly disagree, 1 = disagree, 2 = somewhat disagree, 3 = neutral, 4 = somewhat agree, 5 = agree and 6 = strongly agree. The mean ± SD and median (in brackets) for the query “I could hear LIFU” for AI, aMCC and Sham was: 1.4 ± 1.9 (0); 1.7 ± 2.0 (1) and 1.3 ± 1.8. The mean ± SD and median (in brackets) for the query “I could feel LIFU” for AI, aMCC and Sham was: 1.4 ± 1.7 (1); 1.5 ± 1.6 (1) and 1.2 ± 1.6 (1). The mean ± SD and median (in brackets) for the query “I believe I experienced LIFU” for AI, aMCC and Sham was: 3.5 ± 1.6 (3); 3.4 ± 1.3 (3.5) and 3.1 ± 1.5 (3). No statistical differences were found for any of the queries (all p-adjusted > 0.34). See Supplementary Figure 3.

**Supplementary Figure 3.**
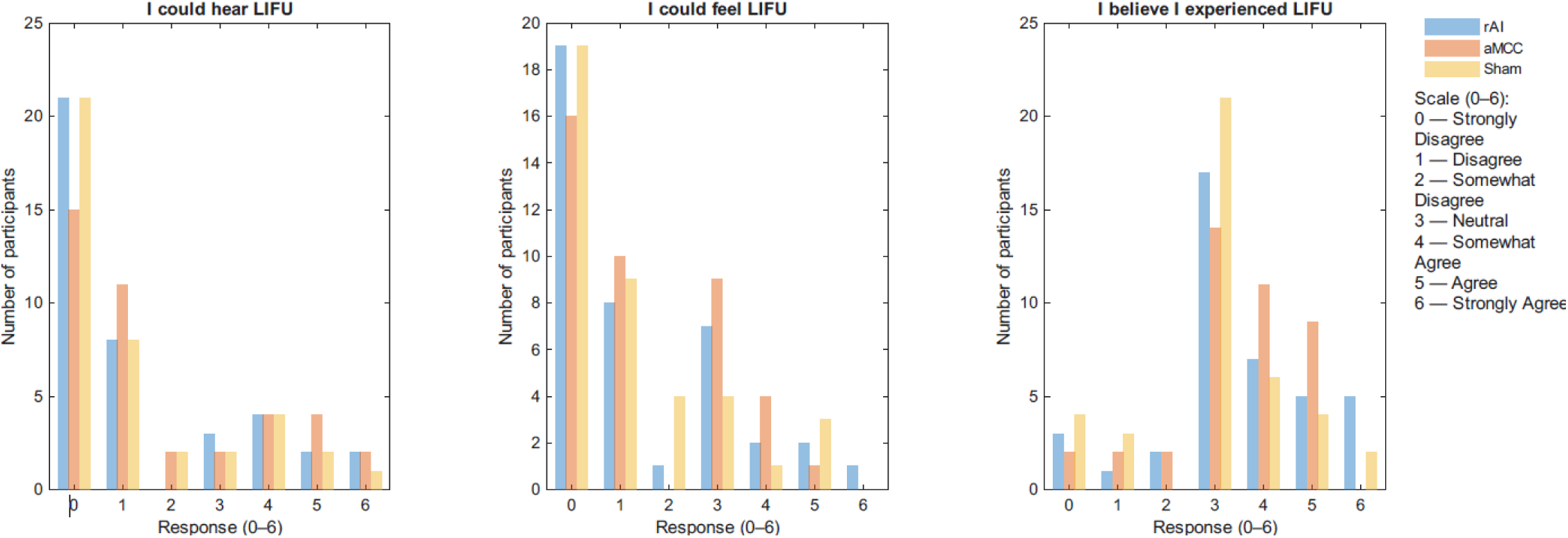

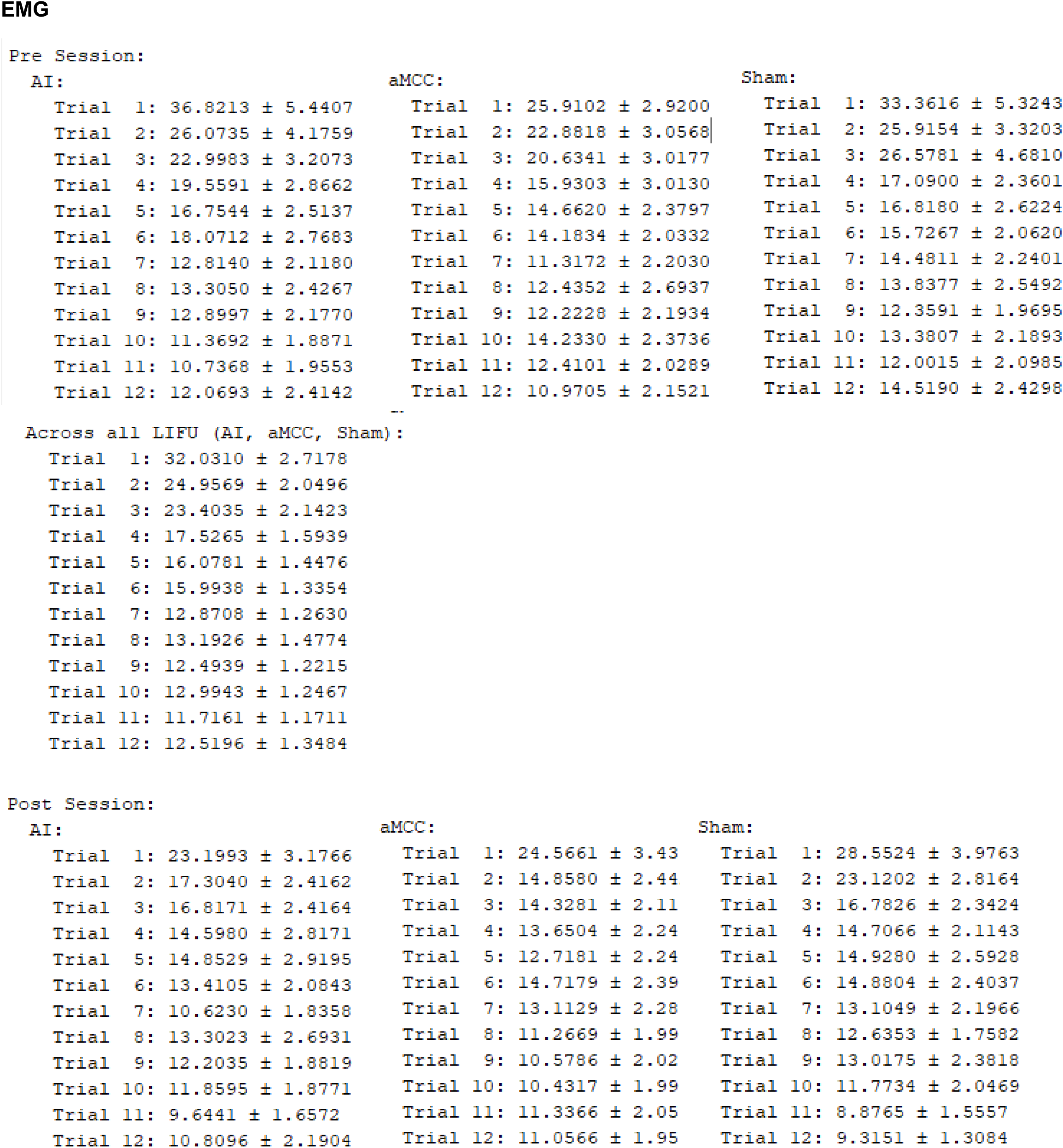
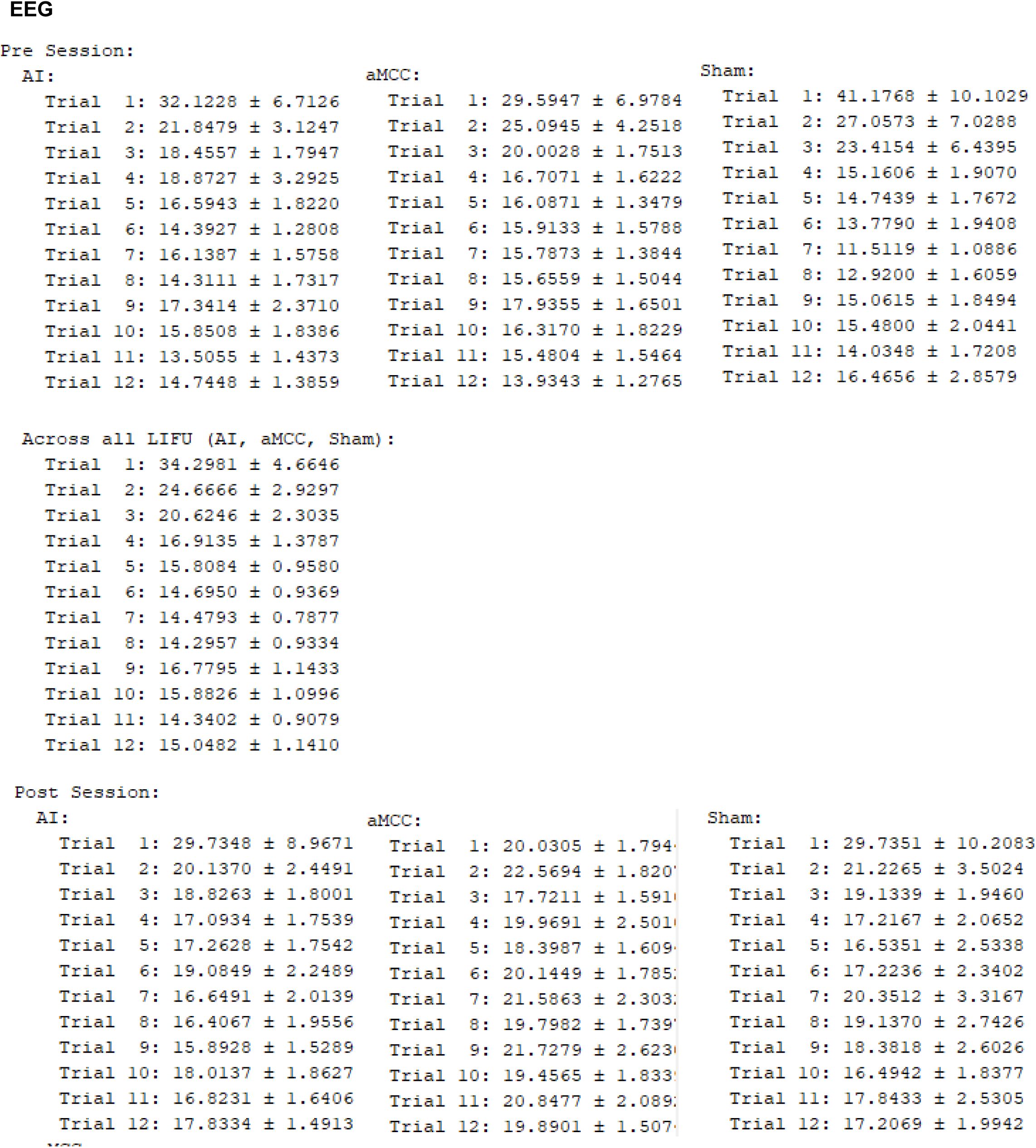
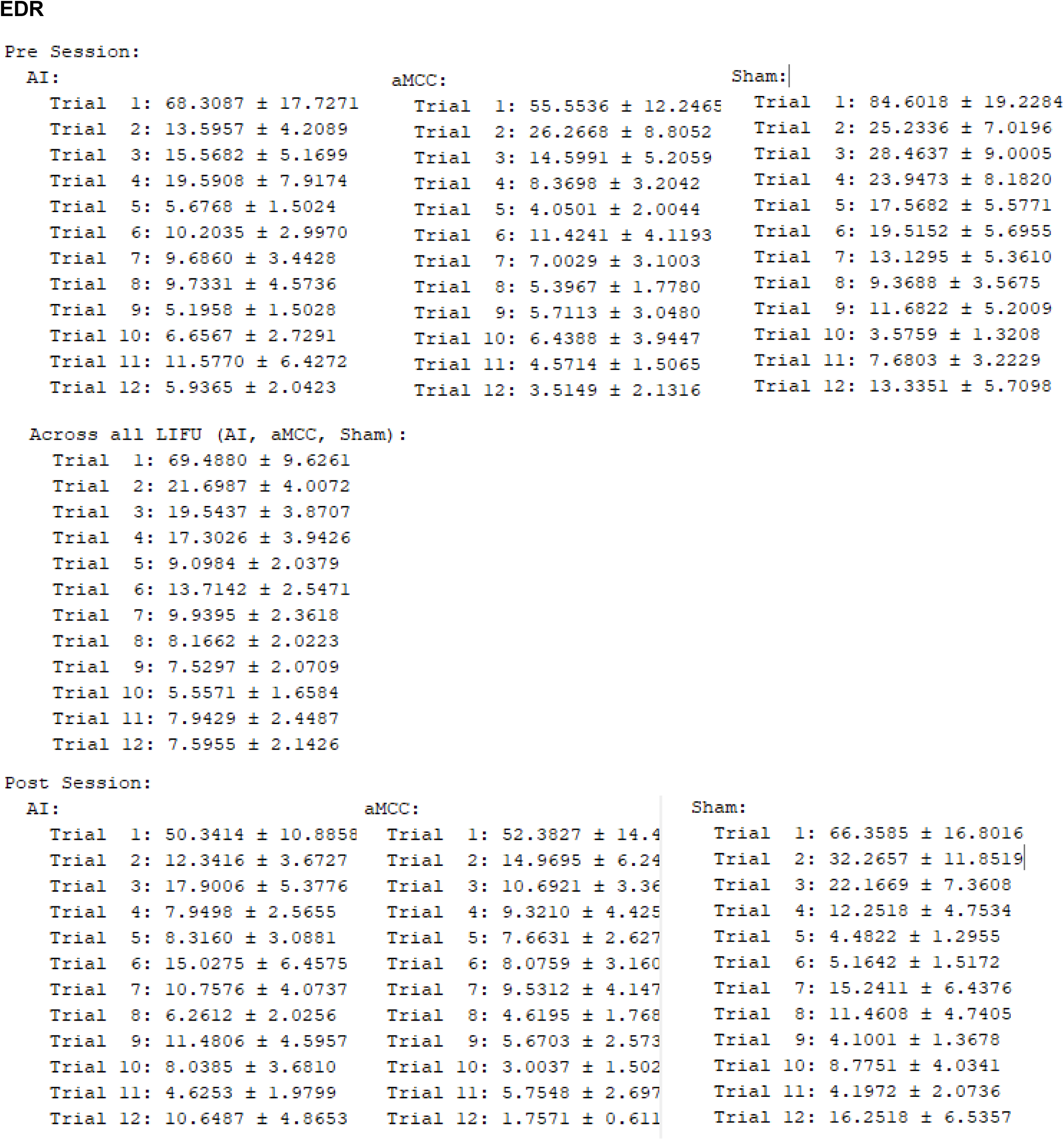

**Supplemental Table 2.**

|  | **EMG** |  |  |
| --- | --- | --- | --- |
|  | *Trial 1* | *Trials 2 - 6* | *Trials 7 - 12* |
| **STAI-T** | $\rho = 0.09$<br>$p = 0.55$ | $\rho = 0.16$<br>$p = 0.34$ | $\rho = 0.10$<br>$p = 0.55$ |
| **STAI-S** | $r = -0.05$<br>$p = 0.62$ | $r = -0.15$<br>$p = 0.19$ | $r = -0.03$<br>$p = 0.78$ |
| **AI vs. Sham** | $t(234) = -1.63$<br>$p = 0.10$ | $t(234) = -0.73$<br>$p = 0.46$ | $t(234) = 0.63$<br>$p = 0.53$ |
| **aMCC vs. Sham** | $t(234) = 0.64$<br>$p = 0.52$ | $t(234) = -0.03$<br>$p = 0.98$ | $t(234) = 0.53$<br>$p = 0.59$ |
|  | **EEG** |  |  |
|  | *Trial 1* | *Trials 2 - 6* | *Trials 7 - 12* |
| **STAI-T** | $\rho = -0.02$<br>$p = 0.90$ | $\rho = 0.09$<br>$p = 0.60$ | $\rho = 0.09$<br>$p = 0.57$ |
| **STAI-S** | $r = -0.11$<br>$p = 0.31$ | $r = 0.02$<br>$p = 0.85$ | $r = -0.05$<br>$p = 0.65$ |
| **AI vs. Sham** | $t(234) = 0.65$<br>$p = 0.52$ | $t(234) = 0.32$<br>$p = 0.75$ | $t(234) = -1.05$<br>$p = 0.29$ |
| **aMCC vs. Sham** | $t(234) = 0.14$<br>$p = 0.89$ | $t(234) = 0.49$<br>$p = 0.62$ | $t(234) = 0.31$<br>$p = 0.75$ |
|  | **EDR** |  |  |
|  | *Trial 1* | *Trials 2 - 6* | *Trials 7 - 12* |
| **STAI-T** | $\rho = -0.13$<br>$p = 0.42$ | $\rho = -0.06$<br>$p = 0.71$ | $\rho = -0.08$<br>$p = 0.61$ |
| **STAI-S** | $r = 0.24$<br>**$p = 0.03$** | $r = -0.024$<br>$p = 0.83$ | $r = -0.08$<br>$p = 0.46$ |
| **AI vs. Sham** | $t(234) = 0.01$<br>$p = 0.99$ | $t(234) = 1.24$<br>$p = 0.23$ | $t(234) = 0.07$<br>$p = 0.94$ |
| **aMCC vs. Sham** | $t(234) = 0.78$<br>$p = 0.43$ | $t(234) = 0.83$<br>$p = 0.41$ | $t(234) = -0.14$<br>$p = 0.88$ |

